# Accessibility and acceptability of a multi-sector collaborative project to address birth inequities: A mixed-methods study

**DOI:** 10.1101/2025.06.05.25329086

**Authors:** Osamuedeme J. Odiase, Alison M. El Ayadi, Malini A. Nijagal, April J. Bell, Kattia Vargas, Prisca C. Diala, Emma Kang, Chinomnso Okorie, Julia Viega, Ntemena Kapula, Garrett Jacobs, KaSelah Crockett, Patience A. Afulani

## Abstract

The Pregnancy Village (PV) was created to deliver cross-sector services at a convenient time and location for Black pregnant and postpartum people. This study examines the accessibility and acceptability of PV among pregnant individuals and their families. We surveyed 116 participants (57 pregnant/postpartum individuals and 59 family members) and conducted 18 semi-structured interviews between July 2021 and June 2022. Accessibility was measured using a 3-item scale and acceptability with a 7-item scale based on the Theoretical Framework of Acceptability; scores were standardized to a 0-100 range. Descriptive, bivariate, and multivariate analyses were conducted, along with framework analysis of qualitative data. Mean accessibility was 76.0 (*SD*=21.0), with mixed qualitative perspectives. Acceptability was high (*M*=91.9, *SD*=14.4), though lower among participants reporting prenatal care discrimination. Qualitative findings supported high acceptability, highlighting PV’s welcoming, supportive atmosphere. Findings suggest PV is accessible and acceptable, with continued model iteration critical for ensuring community priorities and sustainability.

## INTRODUCTION

The United States (U.S.) has the highest maternal mortality rate among high-income countries, with stark inequities in critical perinatal outcomes, including severe maternal morbidity, preterm birth, and infant death (Gunja et al., *Health and Health Care for Women of Reproductive Age*, https://www.commonwealthfund.org/publications/issue-briefs/2022/apr/health-and-health-care-women-reproductive-age). Black women and gender-diverse birthing people (we use ‘individuals’ for brevity) are about three times more likely than White individuals to die from pregnancy-related complications, and their infants are twice as likely to die before the age of one (Hoyert, *Maternal Mortality Rates in the United States, 2022*, https://www.cdc.gov/nchs/data/hestat/maternal-mortality/2022/maternal-mortality-rates-2022.htm; CDC, *Infant Mortality in the United States, 2022: Data from the Period Linked Birth/Infant Death File*, Natl. Vital Stat. Rep. 73, https://www.cdc.gov/nchs/data/nvsr/nvsr73/nvsr73-05.pdf). Even in states like California, where maternal mortality rates are declining—with a ratio of 10.5 deaths per 100,000 live births, (National Center for Health Statistics, *Maternal Mortality by State, 2018–2022*, https://www.cdc.gov/nchs/maternal-mortality/mmr-2018-2022-state-data.pdf) significantly lower than the national average of 22.3 deaths per 100,000 live births—these disparities persist. (Chcf, *Maternity Care in California, 2019: A Bundle of Data*, 46, https://www.chcf.org/wp-content/uploads/2019/11/MaternityCareCAAlmanac2019.pdf; Hoyert, *Maternal Mortality Rates in the United States, 2022*, 2024, https://www.cdc.gov/nchs/data/hestat/maternal-mortality/2022/maternal-mortality-rates-2022.pdf).

The factors driving inequities in maternal and neonatal health are complex, but racism is a key driver, shaping both access to and experiences of high-quality care.^1–3^ For instance, Black and other racially or ethnically minoritized individuals are more likely to have lower incomes and face barriers to receiving high-quality care (Artiga et al., *Racial Disparities in Maternal and Infant Health: An Overview - Issue Brief*, KFF, https://www.kff.org/report-section/racial-disparities-in-maternal-and-infant-health-an-overview-issue-brief/).^2^ While public insurance programs like Medicaid can help bridge the financial gap for service costs, additional challenges such as transportation, childcare, and limited time off work can complicate care access (Artiga et al., *Racial Disparities in Maternal and Infant Health: An Overview - Issue Brief*, KFF, https://www.kff.org/report-section/racial-disparities-in-maternal-and-infant-health-an-overview-issue-brief/).^2^ Discrimination and lack of person-centered care—defined as care that is responsive to and respectful of individuals’ needs, preferences, and values—are also common.^1,4^ Black individuals often report being disrespected, ignored, or not believed during care encounters, reflecting interpersonally mediated racism.^5–8^ These experiences affect the receipt of timely and appropriate care even without logistical barriers.^1,9^ Inequitable healthcare access and experience compound the pervasive stress of everyday discrimination, thus reducing the health and well-being of Black individuals. Interventions addressing equitable access to care and eliminating racism and discrimination in care settings are needed to improve health outcomes, especially in pregnancy, where the needs and timeline are more concentrated.

Several innovative perinatal care programs across the U.S. have sought to improve perinatal outcomes and address birth inequities among Black birthing people through culturally responsive, community-centered approaches. These include California’s Black Infant Health Program, which offers group-based care and social support (*Black Infant Health – Empowering Pregnant and Mothering Black Women*, https://blackinfanthealth.org); Florida’s JJ Way® Clinic, a patient-centered care model that emphasizes equitable care access, provision of dignified and culturally responsive care, and wraparound services (*The JJ Way® - Commonsense Childbirth Inc.*, https://commonsensechildbirth.org/the-jj-way); and Washington, D.C.’s Mamatoto Village, which integrates perinatal care with workforce development and community advocacy (*Mamatoto Village*, https://www.mamatotovillage.org). However, to our knowledge, no cross-sector, community-centered, community-institutional co-led care delivery models exist to address birth inequities in San Francisco (SF) or elsewhere. This gap led our team to embark on a one-year Human-Centered Design (HCD) process in 2017, focused on SF residents facing the most severe inequities, particularly those insured by Medicaid and identifying as Black. Despite SF’s reputation as a progressive city with many equity-focused perinatal programs, participants described excessive practical barriers to care and rampant experiences of interpersonal racism. Referrals between agencies also caused stress because services were fragmented and disconnected.^10^ Our results confirmed extensive evidence that U.S. healthcare delivery systems—built within a culture of white supremacy—often structure care that is difficult to access, ^11,12^ fails to meet individuals’ needs, and undermines the autonomy and dignity of Black pregnant individuals.^1,5,8^

Through the HCD process emerged the “Pregnancy Village” model: a community-institutional collaboration designed to provide a “one-stop shop” of cross-sector services (clinical, city/government, and community-based) in a safe, healing, and uplifting environment, with a focus on earning trust and *comprehensively* improving health and wellness of individuals, families, and communities. Through a formal partnership, community-based organizations, institutions (e.g., health systems and city government agencies), and community members with lived experience would co-design and implement the model together, ensuring it is culturally relevant, responsive to community needs, and driven by community priorities. Key model tenets include the integration of anti-racism principles across all aspects of model design and service delivery and leveraging community-institutional partnerships to ensure effectiveness and sustainability. Additional details on the development and implementation of PV can be found elsewhere.^10,13^

After a three-year planning process, including a partnership with the “Pop-Up Village” event model founders, Designing Justice + Designing Spaces, the first iteration of the Pregnancy Village—the “SF Family & Pregnancy Pop-Up Village” event series (subsequently referred to as the “Pregnancy Village” (PV) for brevity)—was launched.^13^ PV was piloted during 26 monthly events from July 2021 to June 2024.

Given that the sustainability of any health program is predicated on its accessibility and acceptability, we evaluated the accessibility and acceptability of PV. We utilized the theoretical framework of acceptability (TFA).^14^ Accessibility is captured under acceptability in the TFA, which we operationalized by exploring pregnant and postpartum participants and their families’ *perceived affective attitude* towards the intervention, the effort required to engage with PV (*accessibility* and *burden*), their perceptions of intervention *effectiveness*, and their confidence in seeking out resources outside of PV (*self-efficacy*). Our evaluation is situated within our team’s broader mission of continually engaging community members in model iteration to ensure responsiveness to their needs; thus, we focus the current analysis on the most formative phase: the first nine monthly events of PV implementation (July 2021 to June 2022).

## RESULTS

### Participant Characteristics

The analytic sample comprises 116 participants, as four were determined ineligible after a review of demographic data. Fifteen participants completed the survey more than once. There are, thus, 89 unique individuals in the study. Participants’ sociodemographic and reproductive health characteristics are summarized in Table 1. About half (49%) of participants were currently or recently pregnant (within the past year). Nearly half of the participants were either between 25-34 (27%) or 35-44 years of age (22%). Forty-one percent of participants identified as Black or African American, and 36% identified as Latine. Forty-five percent had an educational level beyond high school, and 18% had not completed high school. About two-thirds (64%) of participants were unemployed, and nearly half (47%) were on income assistance. Two-thirds (66%) of participants had public insurance (e.g., Medi-Cal, Medicaid, etc.). Forty-two percent of participants were single, and 32% lived with a romantic partner. Twenty percent of participants had at least one preterm birth, and one-fourth (25%) had a prior pregnancy loss.

**Table 1.** Univariate distribution of sociodemographic characteristics, obstetric history, and care discrimination experiences among pregnant, postpartum, and family participants in the San Francisco Family & Pregnancy Pop-Up Village, San Francisco, CA

| Participant Characteristics | Total (N=116) | Currently pregnant/recently pregnant (n=57) | Family (n=59) |
| --- | --- | --- | --- |
|  | % (N) | % (n) | % (n) |
| *Age* |  |  |  |
| 15-24 | 11.2% (13) | 17.5% (10) | 5.1% (3) |
| 25-34 | 26.7% (31) | 38.6% (22) | 15.3% (9) |
| 35-44 | 21.6% (25) | 33.3% (19) | 10.2% (6) |
| 45 and older | 25.9% (30) | 1.8% (1) | 49.2% (29) |
| Unknown | 14.7% (17) | 8.8% (5) | 20.3% (12) |
| *Gender* |  |  |  |
| Female | 86.2% (100) | 91.2% (52) | 81.4% (48) |
| Male | 5.2% (6) | 0.0% (0) | 10.2% (6) |
| Other/unknown/prefer not to answer | 8.6% (10) | 8.8% (5) | 8.5% (5) |
| *Race/ethnicity* |  |  |  |
| Black | 40.5% (47) | 26.3% (15) | 54.2% (32) |
| Latina/Hispanic | 36.2% (42) | 47.4% (27) | 25.4% (15) |
| Multiracial | 15.5% (18) | 19.3% (11) | 11.9% (7) |
| Other/unknown/prefer not to answer | 7.8% (9) | 7.0% (4) | 8.5% (5) |
| *Language* |  |  |  |
| English | 59.5% (69) | 52.6% (30) | 66.1% (39) |
| Spanish | 30.2% (35) | 38.6% (22) | 22.0% (13) |
| Unknown/other/prefer not to answer | 10.3% (12) | 8.8% (5) | 11.9% (7) |
| *English proficiency* |  |  |  |
| Very well or well | 70.7% (82) | 70.2% (40) | 71.2% (42) |
| With difficulty | 17.2% (20) | 19.3% (11) | 15.3% (9) |
| Unknown/prefer not to answer | 12.1% (14) | 10.5% (6) | 13.6% (8) |
| *Pregnancy status* |  |  |  |
| Family member | 50.9% (59) | 0.0% (0) | 100.0% (59) |
| Pregnant or postpartum | 49.1% (57) | 100.0% (57) | 0.0% (0) |
| *Parity* |  |  |  |
| No births | 22.4% (26) | 26.3% (15) | 18.6% (11) |
| 1-2 births | 43.1% (50) | 47.4% (27) | 39.0% (23) |
| 3 or more births | 24.1% (28) | 24.6% (14) | 23.7% (14) |
| Unknown/not applicable/ prefer not to answer | 10.3% (12) | 1.8% (1) | 18.6% (11) |
| *Prenatal care* |  |  |  |
| No prenatal care during current/recent pregnancy | 3.4% (4) | 7.0% (4) | 0.0% (0) |
| Received prenatal care during current/recent pregnancy | 41.4% (48) | 84.2% (48) | 0.0% (0) |
| Unknown/not applicable/prefer not to answer | 55.2% (64) | 8.8% (5) | 100.0% (59) |
| *History of preterm birth* |  |  |  |
| No preterm births | 69.0% (80) | 80.7% (46) | 57.6% (34) |
| At least 1 preterm birth | 19.8% (23) | 15.8% (9) | 23.7% (14) |
| Unknown/prefer not to answer/not applicable | 11.2% (13) | 3.5% (2) | 18.6% (11) |
| *History of pregnancy loss* |  |  |  |
| No prior pregnancy loss | 59.5% (69) | 57.9% (33) | 61.0% (36) |
| Pregnancy loss | 25.0% (29) | 35.1% (20) | 15.3% (9) |
| Unknown/prefer not to answer/ not applicable | 15.5% (18) | 7.0% (4) | 23.7% (14) |
| *Education* |  |  |  |
| Less than a high school degree | 18.1% (21) | 15.8% (9) | 20.3% (12) |
| High school graduate, GED or equivalent | 25.9% (30) | 29.8% (17) | 22.0% (13) |
| Some college, junior college or vocational school | 27.6% (32) | 21.1% (12) | 33.9% (20) |
| College graduate or higher | 17.2% (20) | 21.1% (12) | 13.6% (8) |
| Unknown/prefer not to answer | 11.2% (13) | 12.3% (7) | 10.2% (6) |
| *Employment* |  |  |  |
| No | 63.8% (74) | 56.1% (32) | 71.2% (42) |
| Yes, Full-time | 13.8% (16) | 19.3% (11) | 8.5% (5) |
| Yes, Part-time | 10.3% (12) | 15.8% (9) | 5.1% (3) |
| Unknown/prefer not to answer | 12.1% (14) | 8.8% (5) | 15.3% (9) |
| *Income assistance* |  |  |  |
| No | 43.1% (50) | 33.3% (19) | 52.5% (31) |
| Yes | 47.4% (55) | 54.4% (31) | 40.7% (24) |
| Unknown/ prefer not to answer | 9.5% (11) | 12.3% (7) | 6.8% (4) |
| *Residence* |  |  |  |
| Bayview-Hunter's Point (San Francisco) | 33.6% (39) | 38.6% (22) | 28.8% (17) |
| Other San Francisco | 44.8% (52) | 43.9% (25) | 45.8% (27) |
| East Bay | 9.5% (11) | 12.3% (7) | 6.8% (4) |
| Other/unknown/prefer not to answer | 12.1% (14) | 5.3% (3) | 18.6% (11) |
| *Housing status* |  |  |  |
| Homeless Shelter | 10.3% (12) | 12.3% (7) | 8.5% (5) |
| Living with someone for free | 4.3% (5) | 5.3% (3) | 3.4% (2) |
| No living place | 0.9% (1) | 0.0% (0) | 1.7% (1) |
| Owens home or apartment | 6.9% (8) | 5.3% (3) | 8.5% (5) |
| Public Housing | 14.7% (17) | 12.3% (7) | 16.9% (10) |
| Renting home or apartment | 50.0% (58) | 52.6% (30) | 47.5% (28) |
| Transitional or supportive housing | 2.6% (3) | 3.5% (2) | 1.7% (1) |
| Other/unknown/prefer not to answer | 10.3% (12) | 8.8% (5) | 11.9% (7) |
| *Social support* |  |  |  |
| Not at all | 12.9% (15) | 7.0% (4) | 18.6% (11) |
| A little | 9.5% (11) | 14.0% (8) | 5.1% (3) |
| Somewhat | 19.0% (22) | 22.8% (13) | 15.3% (9) |
| Yes, definitely | 54.3% (63) | 54.4% (31) | 54.2% (32) |
| Unknown/prefer not to answer | 4.3% (5) | 1.8% (1) | 6.8% (4) |
| *Medical insurance status* |  |  |  |
| Private insurance | 15.5% (18) | 19.3% (11) | 11.9% (7) |
| Public insurance | 66.4% (77) | 66.7% (38) | 66.1% (39) |
| No insurance | 4.3% (5) | 1.8% (1) | 6.8% (4) |
| Unknown/prefer not to answer | 13.8% (16) | 12.3% (7) | 15.3% (9) |
| *Food insecurity (Worried food would run out)* |  |  |  |
| Often true | 11.2% (13) | 8.8% (5) | 13.6% (8) |
| Sometimes true | 24.1% (28) | 33.3% (19) | 15.3% (9) |
| Never true | 48.3% (56) | 36.8% (21) | 59.3% (35) |
| Unknown/prefer not to answer | 16.4% (19) | 21.1% (12) | 11.9% (7) |
| *Food insecurity (Insufficient funds)* |  |  |  |
| Often true | 11.2% (13) | 12.3% (7) | 10.2% (6) |
| Sometimes true | 27.6% (32) | 36.8% (21) | 18.6% (11) |
| Never true | 43.1% (50) | 33.3% (19) | 52.5% (31) |
| Unknown/prefer not to answer | 18.1% (21) | 17.5% (10) | 18.6% (11) |
| *Relationship status* |  |  |  |
| Partnered, living together | 31.9% (37) | 42.1% (24) | 22.0% (13) |
| Partnered, not living together | 13.8% (16) | 17.5% (10) | 10.2% (6) |
| Single | 42.2% (49) | 24.6% (14) | 59.3% (35) |
| Other/unknown/prefer not to answer | 12.1% (14) | 15.8% (9) | 8.5% (5) |
| *Everyday discrimination experience* |  |  |  |
| Never | 17.2% (20) | 19.3% (11) | 15.3% (9) |
| Rarely | 25.9% (30) | 28.1% (16) | 23.7% (14) |
| Sometimes | 36.2% (42) | 35.1% (20) | 37.3% (22) |
| Often | 16.4% (19) | 15.8% (9) | 16.9% (10) |
| Unknown/prefer not to answer | 4.3% (5) | 1.8% (1) | 6.8% (4) |
| *Prenatal care discrimination experience* |  |  |  |
| Never | 42.2% (49) | 31.6% (18) | 52.5% (31) |
| Rarely | 19.0% (22) | 26.3% (15) | 11.9% (7) |
| Sometimes | 22.4% (26) | 35.1% (20) | 10.2% (6) |
| Often | 6.9% (8) | 5.3% (3) | 8.5% (5) |
| Unknown/prefer not to answer | 9.5% (11) | 1.8% (1) | 16.9% (10) |

### Accessibility of PV

The mean accessibility score was 76.0 (SD 21.0) overall, 73.7 (SD 24.4) for the pregnant and postpartum participants, and 78.5 (SD 18.6) for family members (Table 2). The mean accessibility score for Black participants was 78.0 (SD 20.9) compared to 74.6 (SD 22.5) for participants from other racial and ethnic groups.

**Table 2.** Distribution of standardized Accessibility and Acceptability scale scores among pregnant, postpartum, and family participants in the San Francisco Family & Pregnancy Pop-Up Village, San Francisco, CA.

| Scales | Summary statistics | Combined sample | Pregnant and postpartum | Family member | Black participants | Participants from all other racial/ethnic groups |
| --- | --- | --- | --- | --- | --- | --- |
| Accessibility | *N* | 112 | 56 | 56 | 47 | 65 |
|  | *Mean* | 76.0 | 73.2 | 78.9 | 78.0 | 74.6 |
|  | *SD* | 21.8 | 24.3 | 18.7 | 20.9 | 22.5 |
|  | *Minimum Score* | 0.0 | 0.0 | 16.7 | 0.0 | 0.0 |
|  | *Maximum Score* | 100.0 | 100.0 | 100.0 | 100 | 100.0 |
| Acceptability | *N* | 114 | 57 | 57 | 47 | 67 |
|  | *Score* | 91.9 | 90.3 | 93.5 | 94.2 | 90.3 |
|  | *SD* | 14.4 | 15.8 | 12.8 | 12.8 | 15.3 |
|  | *Minimum Score* | 28.6 | 28.6 | 47.6 | 47.6 | 28.6 |
|  | *Maximum Score* | 100.0 | 100.0 | 100.0 | 100 | 100.0 |

Abbreviations: SD: standard deviation

#### Factors associated with accessibility

In bivariate analyses, several factors were associated with perceived accessibility of the PV. Participants who were between the ages of 25 and 34, had limited English proficiency, and worked full-time had significantly lower perceptions of PV accessibility than participants between the ages of 15 and 24, proficient in English, and unemployed, respectively. Also, those who lived in other areas of San Francisco, reported only “somewhat” having social support, had private or employer-sponsored insurance or no medical insurance had lower perceptions of PV accessibility than participants who lived in the Bayview, reported strong social support (“Yes, definitely”), and had public insurance, respectively. Participants who did not disclose their food insecurity status and experienced discrimination during prenatal care encounters on some occasions had significantly lower perceptions of PV accessibility than those who often experienced food insecurity and had no experiences of discrimination during prenatal care encounters, respectively. Participants with some level of college education, who owned a home or apartment, or were single, had significantly higher perceptions of PV accessibility than participants who did not earn a high school diploma, lived in a homeless shelter, or were married or partnered and living together, respectively (Table 3; Supplementary Table 1).

**Table 3.**
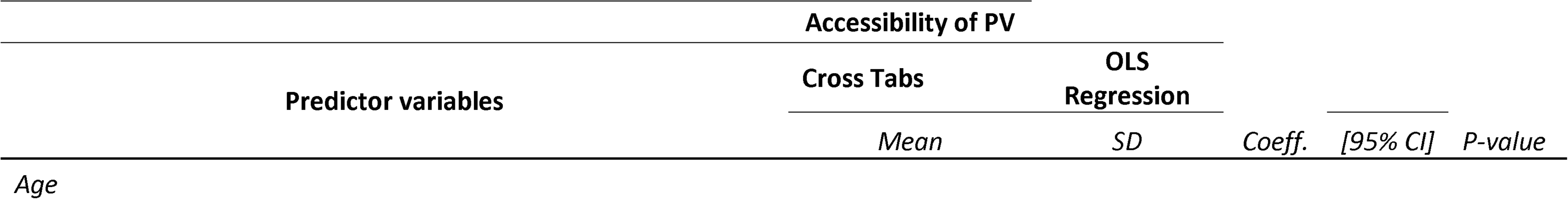
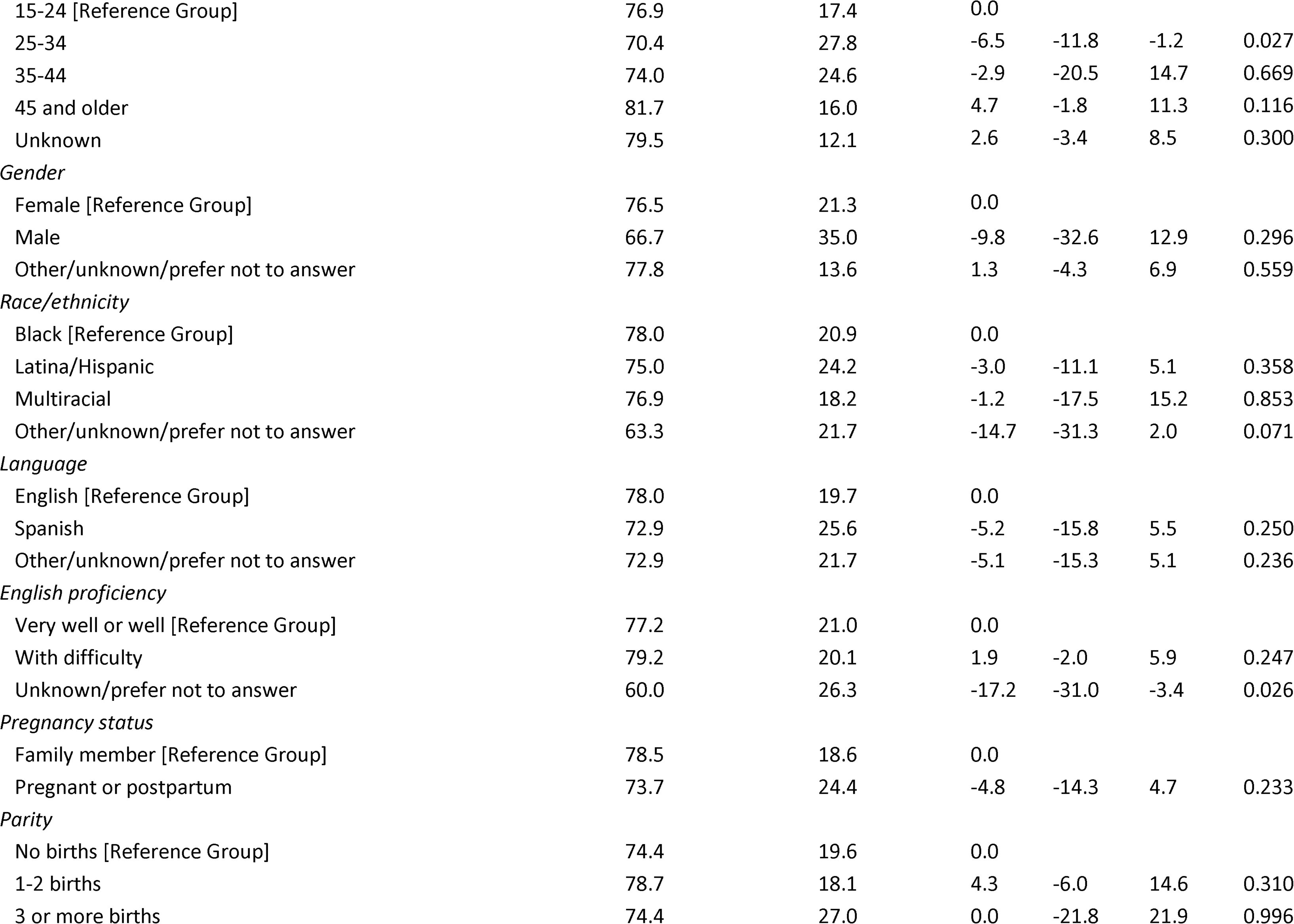
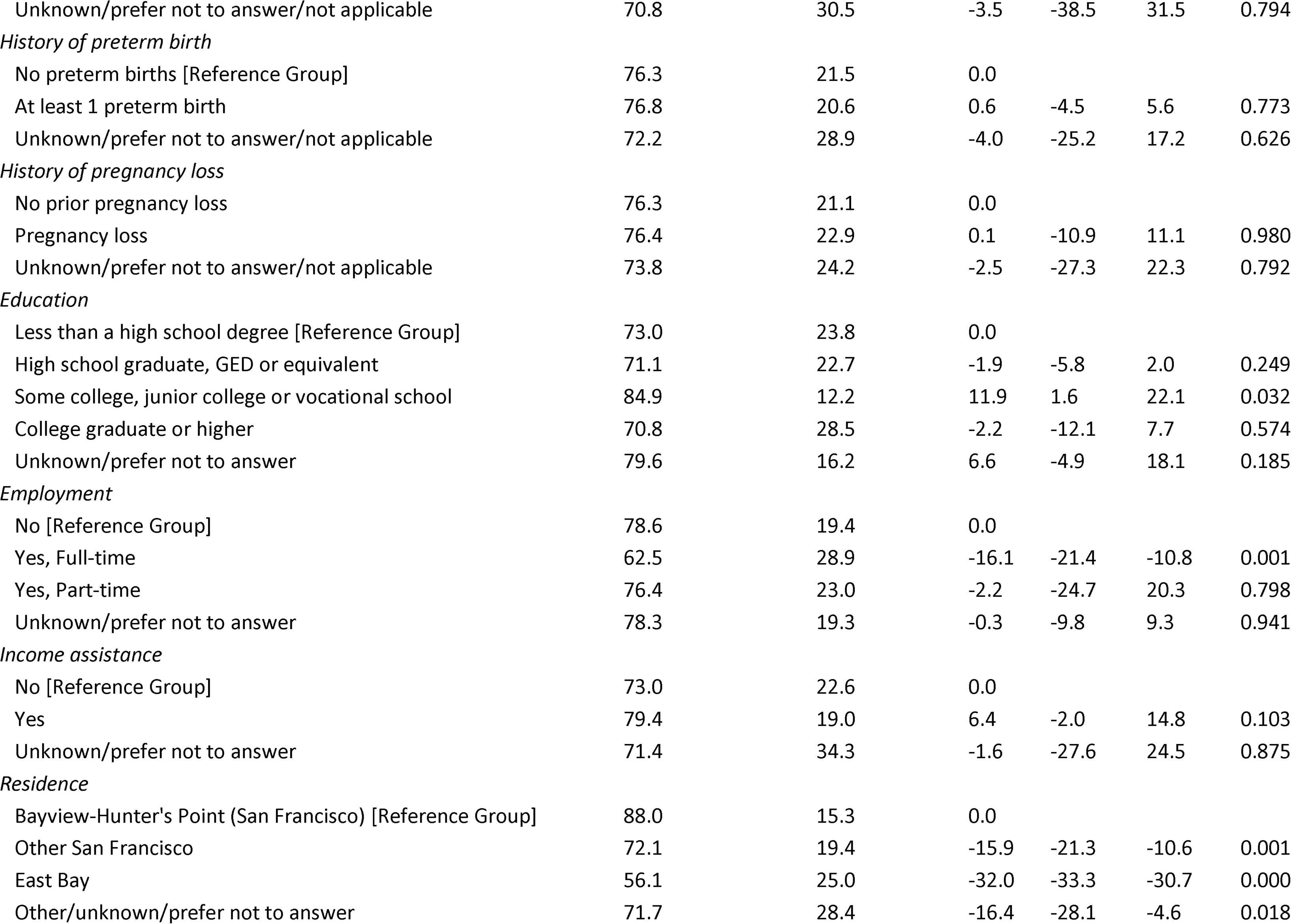
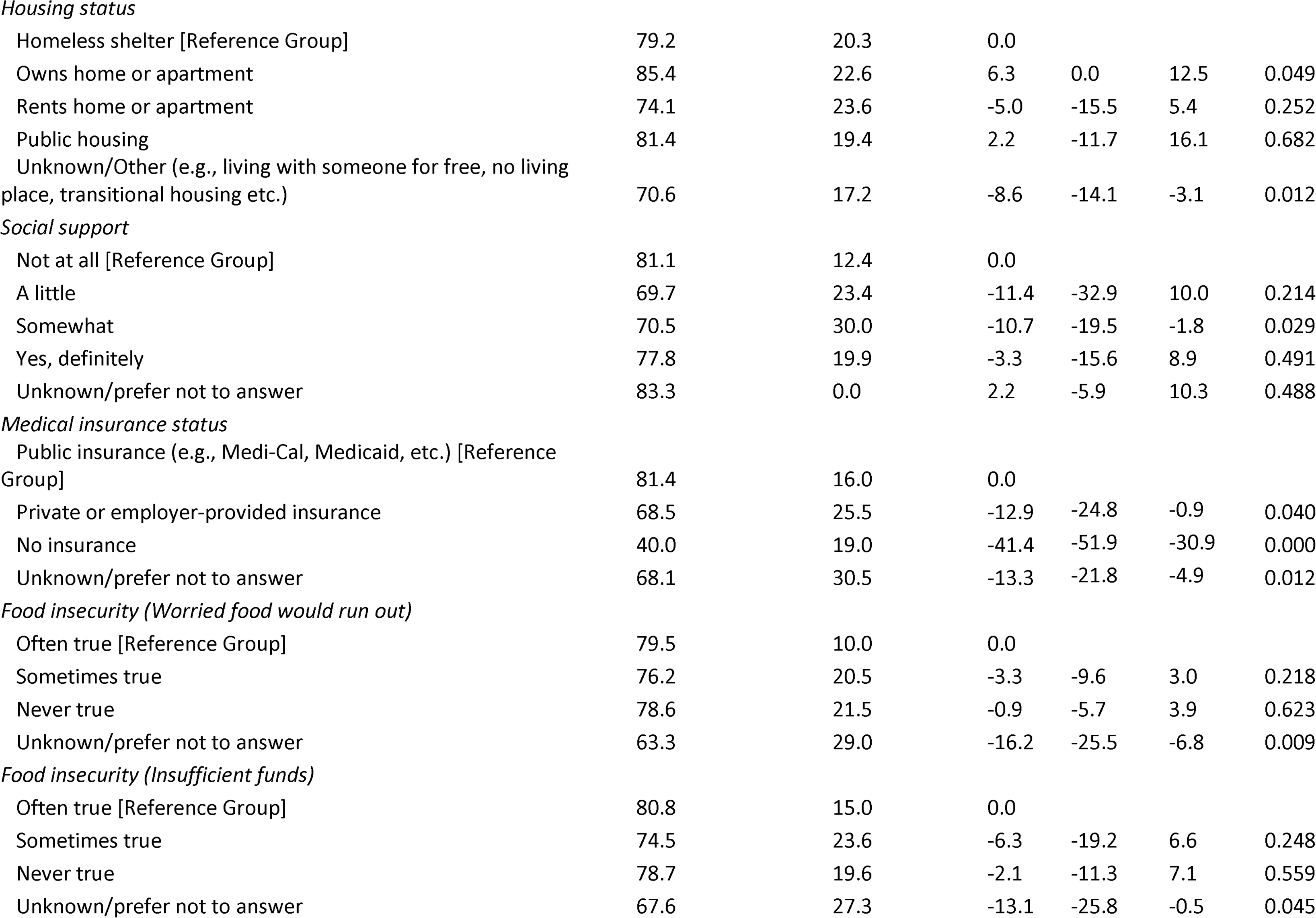
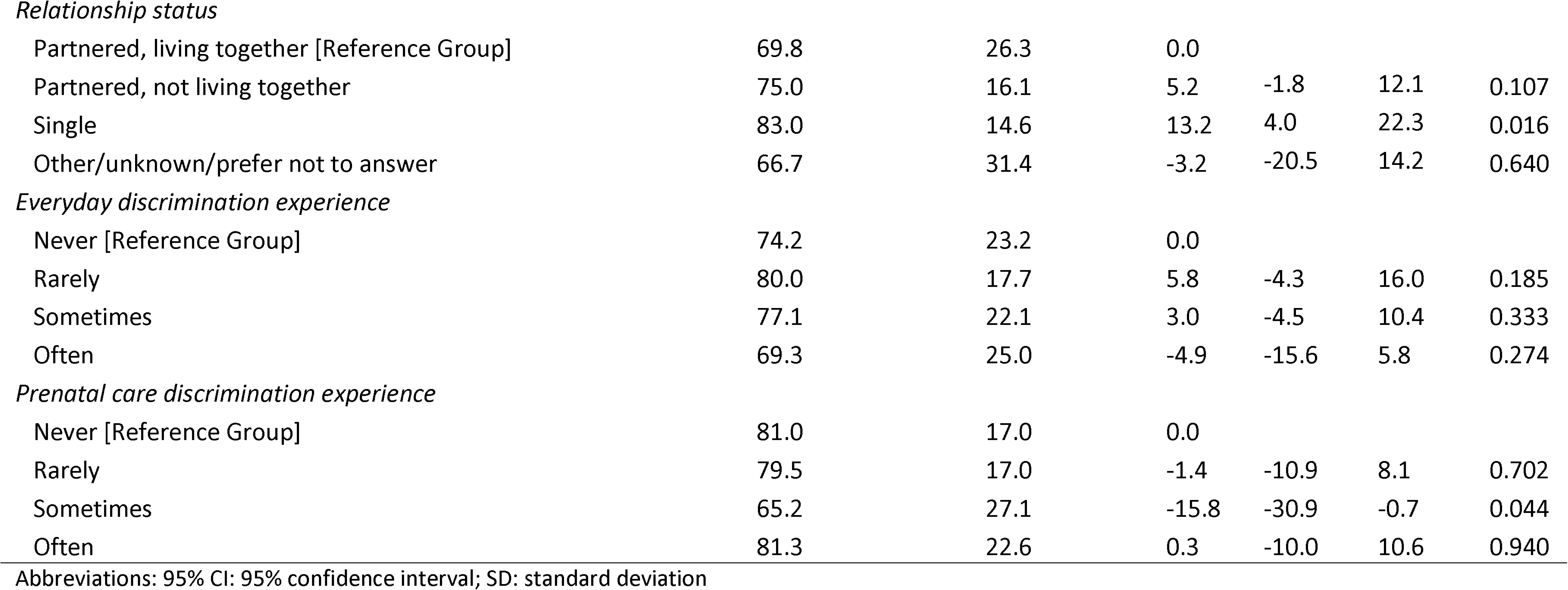
Bivariate statistics of sociodemographic characteristics, obstetric history, and care discrimination experiences for the full sample on the Accessibility score.

| Predictor variables | Accessibility of PV |  |  |  |  |
| --- | --- | --- | --- | --- | --- |
|  | Cross Tabs |  | OLS Regression |  |  |
|  | *Mean* | *SD* | *Coeff.* | *[95% CI]* | *P-value* |
| Age |  |  |  |  |  |

|  |  |  |  |  |  |  |
| --- | --- | --- | --- | --- | --- | --- |
| 15-24 [Reference Group] | 76.9 | 17.4 | 0.0 |  |  |  |
| 25-34 | 70.4 | 27.8 | -6.5 | -11.8 | -1.2 | 0.027 |
| 35-44 | 74.0 | 24.6 | -2.9 | -20.5 | 14.7 | 0.669 |
| 45 and older | 81.7 | 16.0 | 4.7 | -1.8 | 11.3 | 0.116 |
| Unknown | 79.5 | 12.1 | 2.6 | -3.4 | 8.5 | 0.300 |
| *Gender* |  |  |  |  |  |  |
| Female [Reference Group] | 76.5 | 21.3 | 0.0 |  |  |  |
| Male | 66.7 | 35.0 | -9.8 | -32.6 | 12.9 | 0.296 |
| Other/unknown/prefer not to answer | 77.8 | 13.6 | 1.3 | -4.3 | 6.9 | 0.559 |
| *Race/ethnicity* |  |  |  |  |  |  |
| Black [Reference Group] | 78.0 | 20.9 | 0.0 |  |  |  |
| Latina/Hispanic | 75.0 | 24.2 | -3.0 | -11.1 | 5.1 | 0.358 |
| Multiracial | 76.9 | 18.2 | -1.2 | -17.5 | 15.2 | 0.853 |
| Other/unknown/prefer not to answer | 63.3 | 21.7 | -14.7 | -31.3 | 2.0 | 0.071 |
| *Language* |  |  |  |  |  |  |
| English [Reference Group] | 78.0 | 19.7 | 0.0 |  |  |  |
| Spanish | 72.9 | 25.6 | -5.2 | -15.8 | 5.5 | 0.250 |
| Other/unknown/prefer not to answer | 72.9 | 21.7 | -5.1 | -15.3 | 5.1 | 0.236 |
| *English proficiency* |  |  |  |  |  |  |
| Very well or well [Reference Group] | 77.2 | 21.0 | 0.0 |  |  |  |
| With difficulty | 79.2 | 20.1 | 1.9 | -2.0 | 5.9 | 0.247 |
| Unknown/prefer not to answer | 60.0 | 26.3 | -17.2 | -31.0 | -3.4 | 0.026 |
| *Pregnancy status* |  |  |  |  |  |  |
| Family member [Reference Group] | 78.5 | 18.6 | 0.0 |  |  |  |
| Pregnant or postpartum | 73.7 | 24.4 | -4.8 | -14.3 | 4.7 | 0.233 |
| *Parity* |  |  |  |  |  |  |
| No births [Reference Group] | 74.4 | 19.6 | 0.0 |  |  |  |
| 1-2 births | 78.7 | 18.1 | 4.3 | -6.0 | 14.6 | 0.310 |
| 3 or more births | 74.4 | 27.0 | 0.0 | -21.8 | 21.9 | 0.996 |
| Unknown/prefer not to answer/not applicable | 70.8 | 30.5 | -3.5 | -38.5 | 31.5 | 0.794 |
| *History of preterm birth* |  |  |  |  |  |  |
| No preterm births [Reference Group] | 76.3 | 21.5 | 0.0 |  |  |  |
| At least 1 preterm birth | 76.8 | 20.6 | 0.6 | -4.5 | 5.6 | 0.773 |
| Unknown/prefer not to answer/not applicable | 72.2 | 28.9 | -4.0 | -25.2 | 17.2 | 0.626 |
| *History of pregnancy loss* |  |  |  |  |  |  |
| No prior pregnancy loss | 76.3 | 21.1 | 0.0 |  |  |  |
| Pregnancy loss | 76.4 | 22.9 | 0.1 | -10.9 | 11.1 | 0.980 |
| Unknown/prefer not to answer/not applicable | 73.8 | 24.2 | -2.5 | -27.3 | 22.3 | 0.792 |
| *Education* |  |  |  |  |  |  |
| Less than a high school degree [Reference Group] | 73.0 | 23.8 | 0.0 |  |  |  |
| High school graduate, GED or equivalent | 71.1 | 22.7 | -1.9 | -5.8 | 2.0 | 0.249 |
| Some college, junior college or vocational school | 84.9 | 12.2 | 11.9 | 1.6 | 22.1 | 0.032 |
| College graduate or higher | 70.8 | 28.5 | -2.2 | -12.1 | 7.7 | 0.574 |
| Unknown/prefer not to answer | 79.6 | 16.2 | 6.6 | -4.9 | 18.1 | 0.185 |
| *Employment* |  |  |  |  |  |  |
| No [Reference Group] | 78.6 | 19.4 | 0.0 |  |  |  |
| Yes, Full-time | 62.5 | 28.9 | -16.1 | -21.4 | -10.8 | 0.001 |
| Yes, Part-time | 76.4 | 23.0 | -2.2 | -24.7 | 20.3 | 0.798 |
| Unknown/prefer not to answer | 78.3 | 19.3 | -0.3 | -9.8 | 9.3 | 0.941 |
| *Income assistance* |  |  |  |  |  |  |
| No [Reference Group] | 73.0 | 22.6 | 0.0 |  |  |  |
| Yes | 79.4 | 19.0 | 6.4 | -2.0 | 14.8 | 0.103 |
| Unknown/prefer not to answer | 71.4 | 34.3 | -1.6 | -27.6 | 24.5 | 0.875 |
| *Residence* |  |  |  |  |  |  |
| Bayview-Hunter's Point (San Francisco) [Reference Group] | 88.0 | 15.3 | 0.0 |  |  |  |
| Other San Francisco | 72.1 | 19.4 | -15.9 | -21.3 | -10.6 | 0.001 |
| East Bay | 56.1 | 25.0 | -32.0 | -33.3 | -30.7 | 0.000 |
| Other/unknown/prefer not to answer | 71.7 | 28.4 | -16.4 | -28.1 | -4.6 | 0.018 |

*Housing status*
|  |  |  |  |  |  |  |
| --- | --- | --- | --- | --- | --- | --- |
| Homeless shelter [Reference Group] | 79.2 | 20.3 | 0.0 |  |  |  |
| Owns home or apartment | 85.4 | 22.6 | 6.3 | 0.0 | 12.5 | 0.049 |
| Rents home or apartment | 74.1 | 23.6 | -5.0 | -15.5 | 5.4 | 0.252 |
| Public housing | 81.4 | 19.4 | 2.2 | -11.7 | 16.1 | 0.682 |
| Unknown/Other (e.g., living with someone for free, no living place, transitional housing etc.) | 70.6 | 17.2 | -8.6 | -14.1 | -3.1 | 0.012 |

*Social support*
|  |  |  |  |  |  |  |
| --- | --- | --- | --- | --- | --- | --- |
| Not at all [Reference Group] | 81.1 | 12.4 | 0.0 |  |  |  |
| A little | 69.7 | 23.4 | -11.4 | -32.9 | 10.0 | 0.214 |
| Somewhat | 70.5 | 30.0 | -10.7 | -19.5 | -1.8 | 0.029 |
| Yes, definitely | 77.8 | 19.9 | -3.3 | -15.6 | 8.9 | 0.491 |
| Unknown/prefer not to answer | 83.3 | 0.0 | 2.2 | -5.9 | 10.3 | 0.488 |

*Medical insurance status*
|  |  |  |  |  |  |  |
| --- | --- | --- | --- | --- | --- | --- |
| Public insurance (e.g., Medi-Cal, Medicaid, etc.) [Reference Group] | 81.4 | 16.0 | 0.0 |  |  |  |
| Private or employer-provided insurance | 68.5 | 25.5 | -12.9 | -24.8 | -0.9 | 0.040 |
| No insurance | 40.0 | 19.0 | -41.4 | -51.9 | -30.9 | 0.000 |
| Unknown/prefer not to answer | 68.1 | 30.5 | -13.3 | -21.8 | -4.9 | 0.012 |

*Food insecurity (Worried food would run out)*
|  |  |  |  |  |  |  |
| --- | --- | --- | --- | --- | --- | --- |
| Often true [Reference Group] | 79.5 | 10.0 | 0.0 |  |  |  |
| Sometimes true | 76.2 | 20.5 | -3.3 | -9.6 | 3.0 | 0.218 |
| Never true | 78.6 | 21.5 | -0.9 | -5.7 | 3.9 | 0.623 |
| Unknown/prefer not to answer | 63.3 | 29.0 | -16.2 | -25.5 | -6.8 | 0.009 |

*Food insecurity (Insufficient funds)*
|  |  |  |  |  |  |  |
| --- | --- | --- | --- | --- | --- | --- |
| Often true [Reference Group] | 80.8 | 15.0 | 0.0 |  |  |  |
| Sometimes true | 74.5 | 23.6 | -6.3 | -19.2 | 6.6 | 0.248 |
| Never true | 78.7 | 19.6 | -2.1 | -11.3 | 7.1 | 0.559 |
| Unknown/prefer not to answer | 67.6 | 27.3 | -13.1 | -25.8 | -0.5 | 0.045 |
| *Relationship status* |  |  |  |  |  |  |
| Partnered, living together [Reference Group] | 69.8 | 26.3 | 0.0 |  |  |  |
| Partnered, not living together | 75.0 | 16.1 | 5.2 | -1.8 | 12.1 | 0.107 |
| Single | 83.0 | 14.6 | 13.2 | 4.0 | 22.3 | 0.016 |
| Other/unknown/prefer not to answer | 66.7 | 31.4 | -3.2 | -20.5 | 14.2 | 0.640 |
| *Everyday discrimination experience* |  |  |  |  |  |  |
| Never [Reference Group] | 74.2 | 23.2 | 0.0 |  |  |  |
| Rarely | 80.0 | 17.7 | 5.8 | -4.3 | 16.0 | 0.185 |
| Sometimes | 77.1 | 22.1 | 3.0 | -4.5 | 10.4 | 0.333 |
| Often | 69.3 | 25.0 | -4.9 | -15.6 | 5.8 | 0.274 |
| *Prenatal care discrimination experience* |  |  |  |  |  |  |
| Never [Reference Group] | 81.0 | 17.0 | 0.0 |  |  |  |
| Rarely | 79.5 | 17.0 | -1.4 | -10.9 | 8.1 | 0.702 |
| Sometimes | 65.2 | 27.1 | -15.8 | -30.9 | -0.7 | 0.044 |
| Often | 81.3 | 22.6 | 0.3 | -10.0 | 10.6 | 0.940 |

Abbreviations: 95% CI: 95% confidence interval; SD: standard deviation

In the final multivariate model (Table 4), accessibility was significantly associated with English proficiency level, residence, medical insurance status, and experiences of discrimination during prenatal care encounters. Participants who preferred not to disclose their English proficiency level scored, on average, 23.8 points lower than participants who reported being proficient in English (95% CI −40.4, −7.2). Participants who lived in other areas of San Francisco, on average, scored 16.6 points lower than participants who lived in the Bayview (95% CI −24.7, −8.6). Participants who did not have any medical insurance scored, on average, 31.5 points lower than those who had public insurance (95% CI −52.0, −11.0). Participants who experienced discrimination during prenatal care encounters on some occasions scored, on average, 11.0 points lower than those who did not experience discrimination (95%CI: −20.8, −1.2).

**Table 4.**
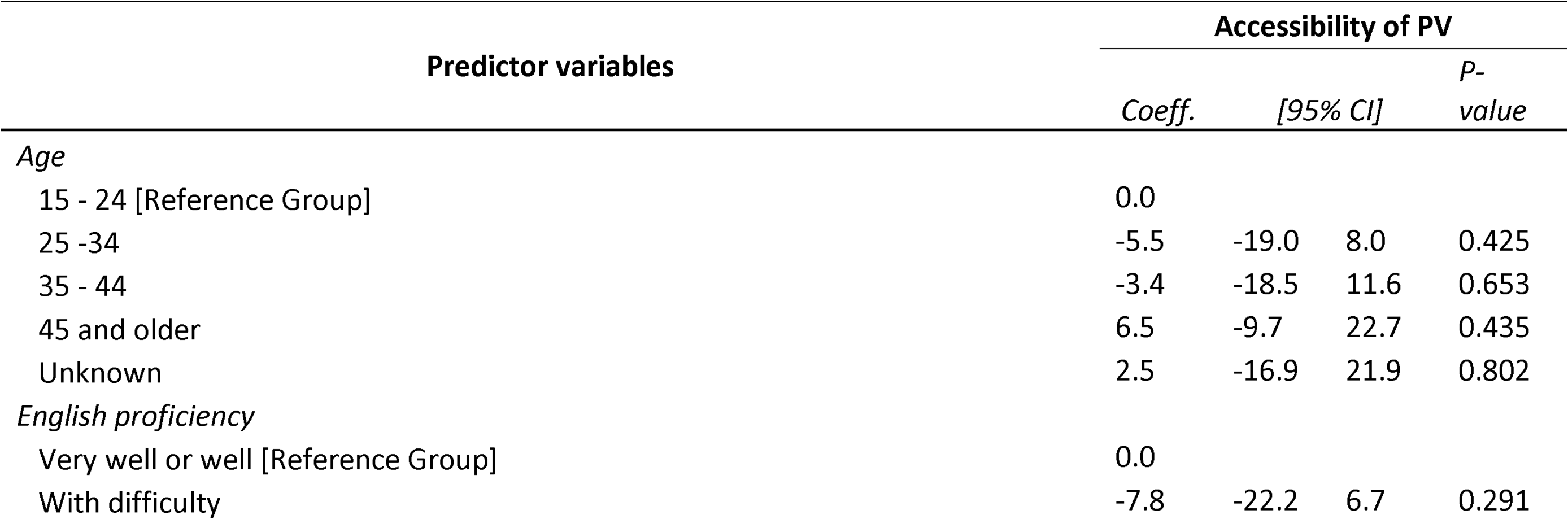
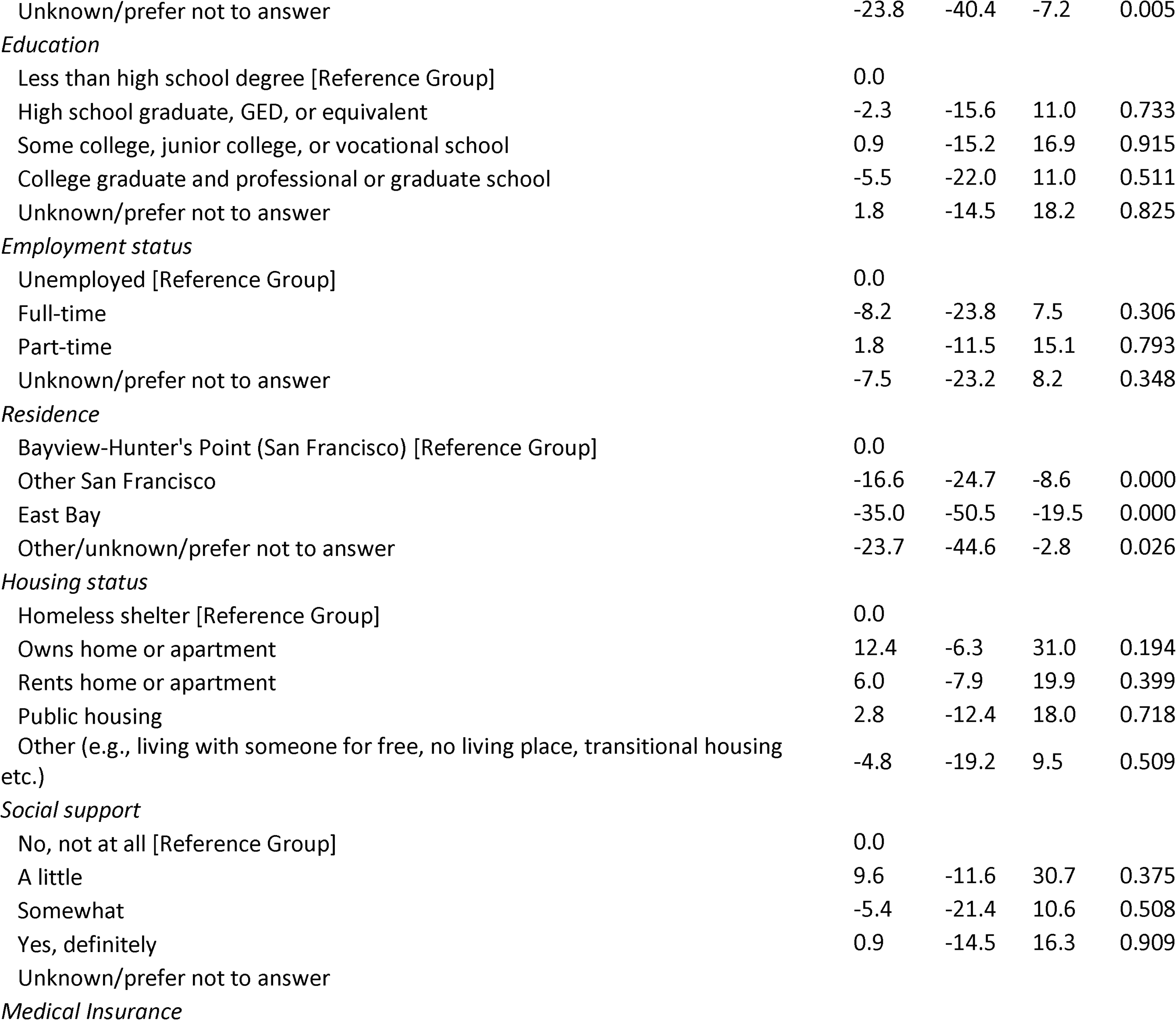
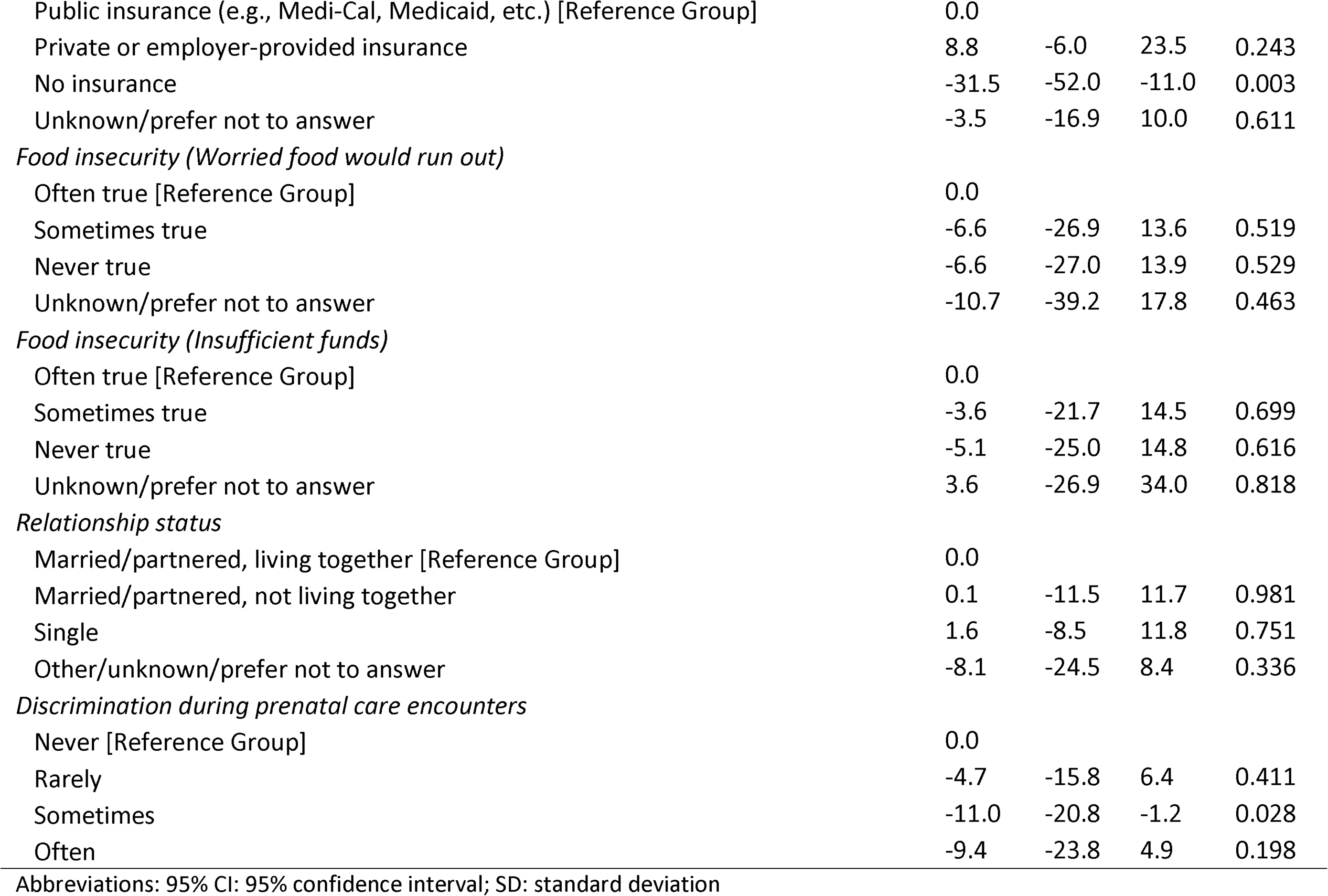
Multivariate mixed-effect linear regression model of select predictors on the Accessibility score.

| Predictor variables | Accessibility of PV |  |  |  |
| --- | --- | --- | --- | --- |
|  | Coeff. | [95% CI] |  | P-value |
| Age |  |  |  |  |
| 15 - 24 [Reference Group] | 0.0 |  |  |  |
| 25 -34 | -5.5 | -19.0 | 8.0 | 0.425 |
| 35 - 44 | -3.4 | -18.5 | 11.6 | 0.653 |
| 45 and older | 6.5 | -9.7 | 22.7 | 0.435 |
| Unknown | 2.5 | -16.9 | 21.9 | 0.802 |
| English proficiency |  |  |  |  |
| Very well or well [Reference Group] | 0.0 |  |  |  |
| With difficulty | -7.8 | -22.2 | 6.7 | 0.291 |
| Unknown/prefer not to answer | -23.8 | -40.4 | -7.2 | 0.005 |
| *Education* |  |  |  |  |
| Less than high school degree [Reference Group] | 0.0 |  |  |  |
| High school graduate, GED, or equivalent | -2.3 | -15.6 | 11.0 | 0.733 |
| Some college, junior college, or vocational school | 0.9 | -15.2 | 16.9 | 0.915 |
| College graduate and professional or graduate school | -5.5 | -22.0 | 11.0 | 0.511 |
| Unknown/prefer not to answer | 1.8 | -14.5 | 18.2 | 0.825 |
| *Employment status* |  |  |  |  |
| Unemployed [Reference Group] | 0.0 |  |  |  |
| Full-time | -8.2 | -23.8 | 7.5 | 0.306 |
| Part-time | 1.8 | -11.5 | 15.1 | 0.793 |
| Unknown/prefer not to answer | -7.5 | -23.2 | 8.2 | 0.348 |
| *Residence* |  |  |  |  |
| Bayview-Hunter's Point (San Francisco) [Reference Group] | 0.0 |  |  |  |
| Other San Francisco | -16.6 | -24.7 | -8.6 | 0.000 |
| East Bay | -35.0 | -50.5 | -19.5 | 0.000 |
| Other/unknown/prefer not to answer | -23.7 | -44.6 | -2.8 | 0.026 |
| *Housing status* |  |  |  |  |
| Homeless shelter [Reference Group] | 0.0 |  |  |  |
| Owns home or apartment | 12.4 | -6.3 | 31.0 | 0.194 |
| Rents home or apartment | 6.0 | -7.9 | 19.9 | 0.399 |
| Public housing | 2.8 | -12.4 | 18.0 | 0.718 |
| Other (e.g., living with someone for free, no living place, transitional housing etc.) | -4.8 | -19.2 | 9.5 | 0.509 |
| *Social support* |  |  |  |  |
| No, not at all [Reference Group] | 0.0 |  |  |  |
| A little | 9.6 | -11.6 | 30.7 | 0.375 |
| Somewhat | -5.4 | -21.4 | 10.6 | 0.508 |
| Yes, definitely | 0.9 | -14.5 | 16.3 | 0.909 |
| Unknown/prefer not to answer |  |  |  |  |
| *Medical Insurance* |  |  |  |  |
| Public insurance (e.g., Medi-Cal, Medicaid, etc.) [Reference Group] | 0.0 |  |  |  |
| Private or employer-provided insurance | 8.8 | -6.0 | 23.5 | 0.243 |
| No insurance | -31.5 | -52.0 | -11.0 | 0.003 |
| Unknown/prefer not to answer | -3.5 | -16.9 | 10.0 | 0.611 |
| *Food insecurity (Worried food would run out)* |  |  |  |  |
| Often true [Reference Group] | 0.0 |  |  |  |
| Sometimes true | -6.6 | -26.9 | 13.6 | 0.519 |
| Never true | -6.6 | -27.0 | 13.9 | 0.529 |
| Unknown/prefer not to answer | -10.7 | -39.2 | 17.8 | 0.463 |
| *Food insecurity (Insufficient funds)* |  |  |  |  |
| Often true [Reference Group] | 0.0 |  |  |  |
| Sometimes true | -3.6 | -21.7 | 14.5 | 0.699 |
| Never true | -5.1 | -25.0 | 14.8 | 0.616 |
| Unknown/prefer not to answer | 3.6 | -26.9 | 34.0 | 0.818 |
| *Relationship status* |  |  |  |  |
| Married/partnered, living together [Reference Group] | 0.0 |  |  |  |
| Married/partnered, not living together | 0.1 | -11.5 | 11.7 | 0.981 |
| Single | 1.6 | -8.5 | 11.8 | 0.751 |
| Other/unknown/prefer not to answer | -8.1 | -24.5 | 8.4 | 0.336 |
| *Discrimination during prenatal care encounters* |  |  |  |  |
| Never [Reference Group] | 0.0 |  |  |  |
| Rarely | -4.7 | -15.8 | 6.4 | 0.411 |
| Sometimes | -11.0 | -20.8 | -1.2 | 0.028 |
| Often | -9.4 | -23.8 | 4.9 | 0.198 |

Abbreviations: 95% CI: 95% confidence interval; SD: standard deviation

Participants’ responses from the in-depth interviews supported the quantitative data on accessibility. While most reported minimal burden of accessing PV services, some reported barriers, including lack of language-concordant navigation services for Spanish-speaking participants and provider follow-up.

#### Facilitators of accessibility

Participants cited several factors that made accessing services easier, including provider responsiveness, short wait times, the physical infrastructure of PV events, and PV’s central location and proximity to their residence. Further, participants emphasized the importance of providers’ willingness to meet them where they were and to work to meet their needs, as one Multiracial pregnant participant explained:

> *“Because you can go up to them, but 9 times out of 10, they are going to come up to you and ask you, “Do you need some of these services?” or “Try this service out.” Because I never got any acupuncture before and that was a new experience for me. But she reached out to me. I didn’t – it wasn’t like I was going over there, so yes.”* —Multiracial pregnant participant less than 30 years old

Other participants noted that services at PV were easier to access due to the short wait times, as participants stated that they typically waited a few minutes before they engaged with providers:

> *“And it wasn’t like a long line. It was just like if I walked to the table; I would be like the first person or like the third. But it wasn’t like a long wait. So, it was like I got to visit the majority of all the programs.”* —Multiracial pregnant participant, between 30 and 44 years old

Participants noted that PV’s central location and proximity to their residence made it convenient for them to walk or take public transportation to reach PV:

> *“You’re [PV] just right off 3rd and McKinnon. That’s just half a block I have to go. Usually, them blocks are long. So, 104 and up there – they [are] long. But it was just a little hop, skip, and a jump, and I was right there. It’s easier for – you didn’t have to walk to catch the bus, or the T-train, the 24, the 54. All of it is right there. Even the 15 comes down that way. So, it’s very easy to get there.”* —Black family member, 45 years old or above

#### Barriers to accessibility

While participants generally reported a low burden in accessing and receiving services at PV events, some discussed challenges related to the physical layout of the space, inadequate service navigation support, overwhelming amount of information, and lack of provider follow-up. Further, participants noted that the difficulty in learning about available services stemmed from a lack of navigators to guide them through the services:

> *“I thought it was going to be like a meeting with […] somebody who could give you details [like a prenatal group], and then I didn’t see anybody, so I was confused. And nobody explained it to us until someone told me, ‘You can go there and get some information.’”* —Latine pregnant participant, between 30 and 44 years old

A few participants noted that lack of follow-up from providers after the event made it difficult to access the services they needed, as this participant recounted their experience:

> *“Well, with [X organization], my experience was good up until they didn’t call me back. They didn’t follow up. So, they were not accountable for their word. So, I felt like [having connected with them] was pointless because … getting that interaction with them and setting that all up with a doula was like the main focus of why I wanted to go. And that’s why my doctor encouraged me to go.”* —Multiracial pregnant participant, less than 30 years old

### Acceptability of the PV

The mean acceptability score was 91.9 (*SD* 14.4) overall, 90.5 (*SD* 15.7) for pregnant and postpartum participants, and 93.4 (*SD* 12.9) for family member participants (Table 2). The mean acceptability score for Black participants was 94.2 (12.8) compared to 90.3 (15.3) for participants from other racial and ethnic groups.

#### Factors associated with acceptability

In bivariate analyses, several factors were associated with perceived acceptability of the PV. Participants who spoke Spanish, had limited English proficiency, and had at least one preterm birth had significantly lower perceptions of PV acceptability than participants who spoke English, were proficient in English, and did not report a history of preterm birth, respectively. Those who had no medical insurance, did not disclose their food insecurity status, and experienced discrimination during prenatal care encounters on some occasions had significantly lower perceptions of PV acceptability than participants who had public insurance, had no food insecurity, and had never been discriminated against during prenatal care encounters, respectively. Participants who worked part-time, owned a home or apartment, and did not experience food insecurity had more positive perceptions of PV acceptability than those who were unemployed, lived in a homeless shelter, and often experienced food insecurity, respectively (Table 5; Supplementary Table 2).

**Table 5.**
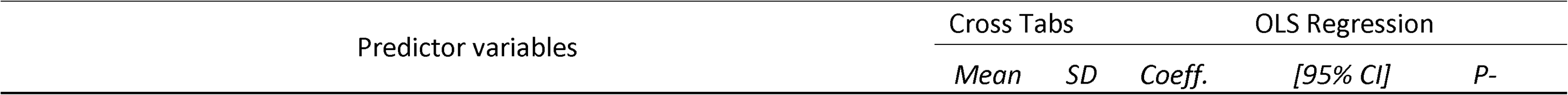
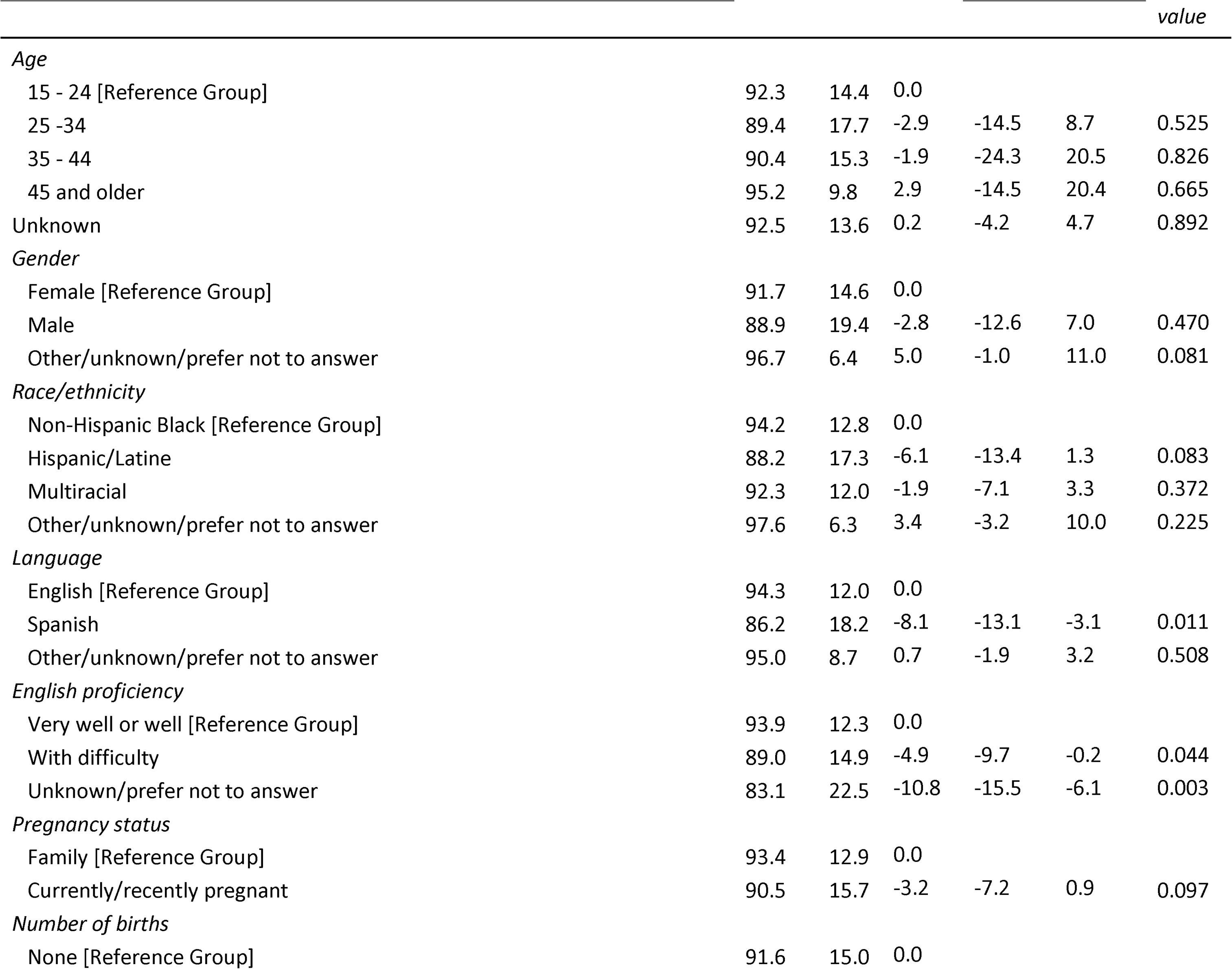
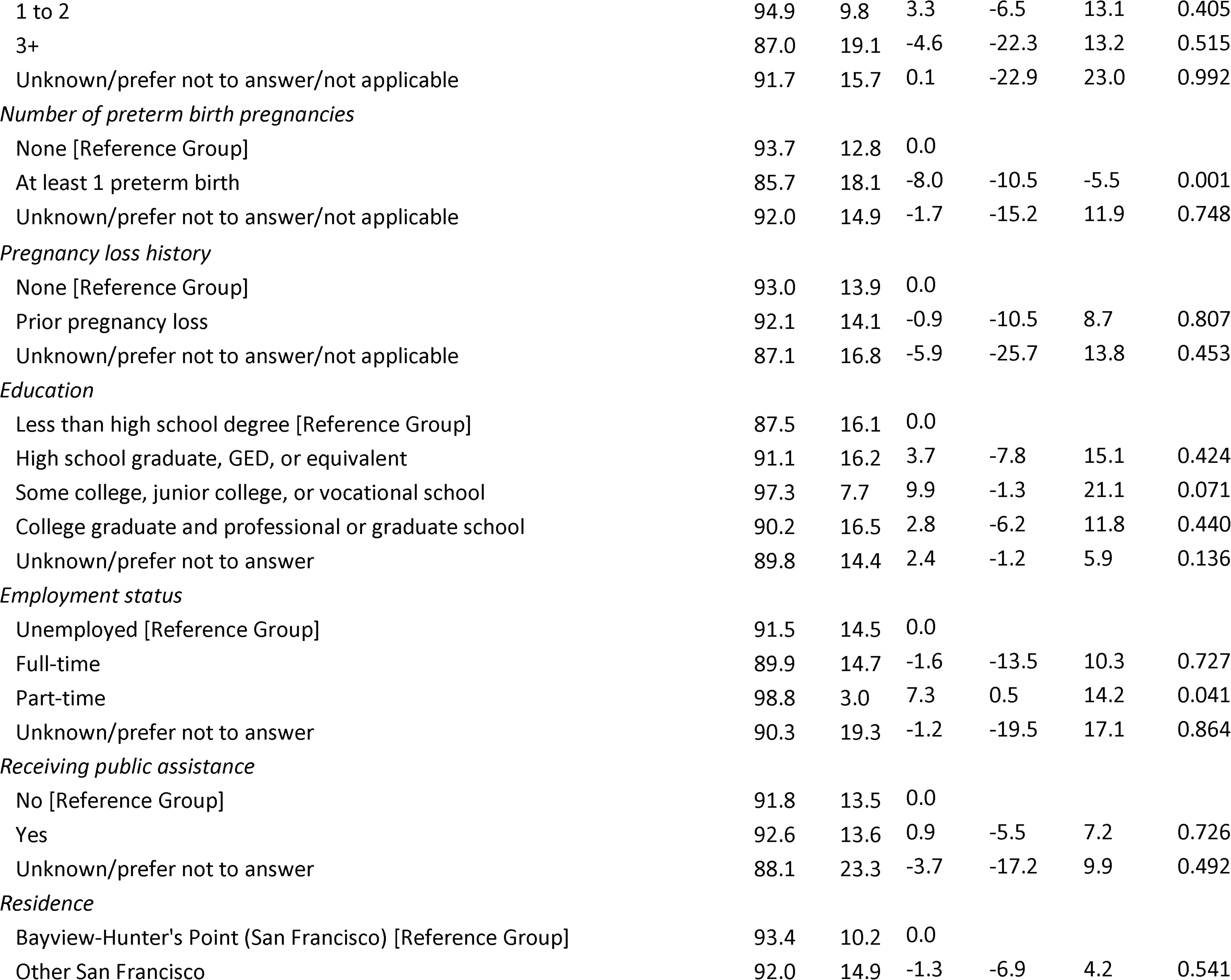
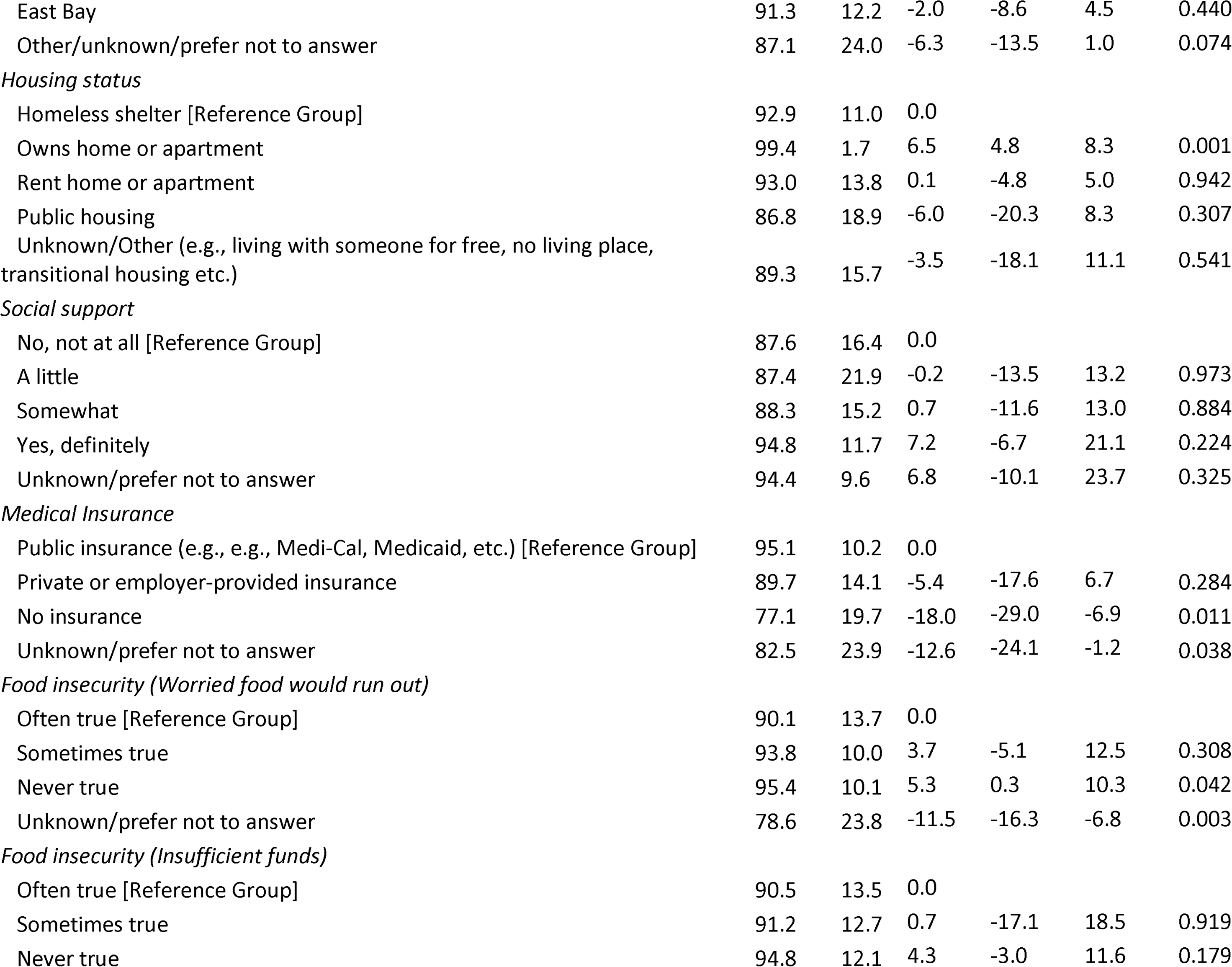
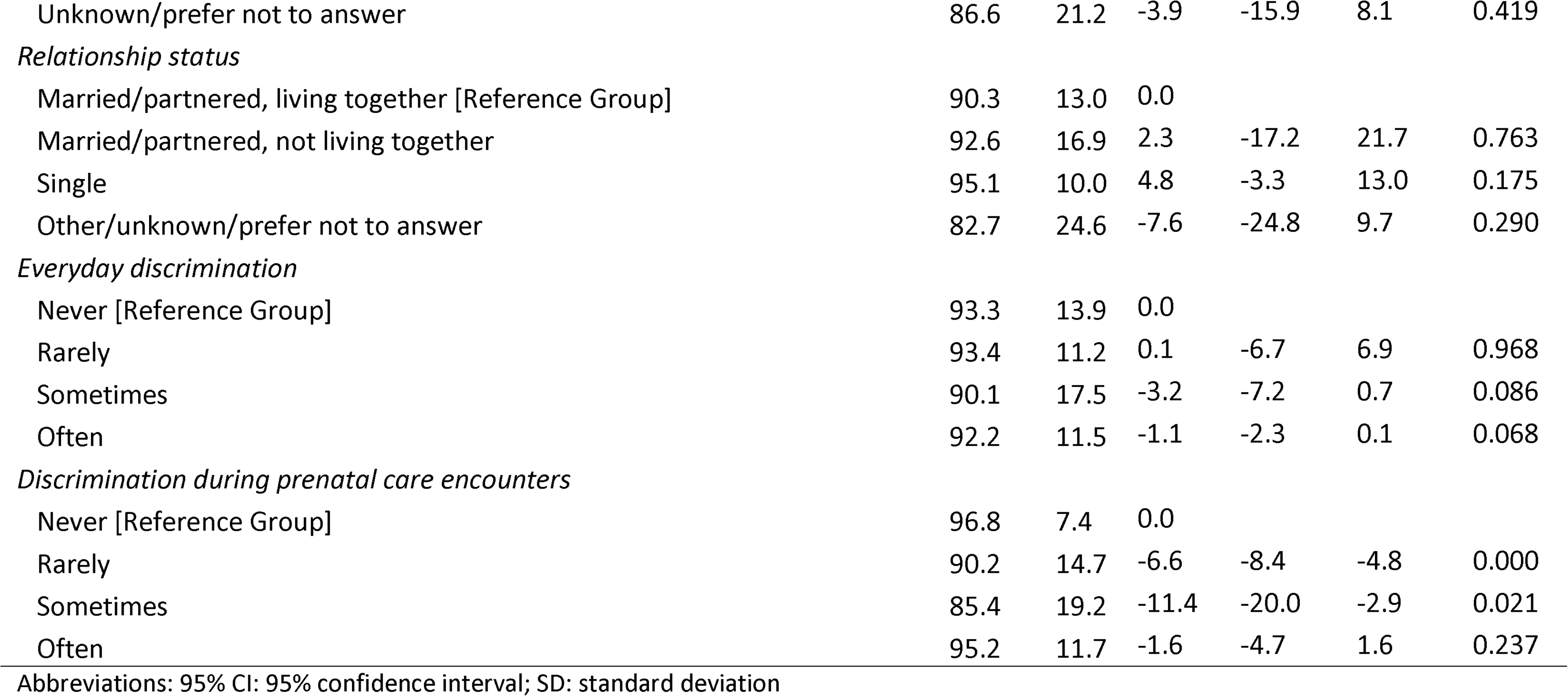
Bivariate statistics of sociodemographic characteristics, obstetric history, and care discrimination experiences for the full sample on the Acceptability score.

|  |  |  |  |  |  | *value* |
| --- | --- | --- | --- | --- | --- | --- |
| *Age* |  |  |  |  |  |  |
| 15 - 24 [Reference Group] | 92.3 | 14.4 | 0.0 |  |  |  |
| 25 -34 | 89.4 | 17.7 | -2.9 | -14.5 | 8.7 | 0.525 |
| 35 - 44 | 90.4 | 15.3 | -1.9 | -24.3 | 20.5 | 0.826 |
| 45 and older | 95.2 | 9.8 | 2.9 | -14.5 | 20.4 | 0.665 |
| Unknown | 92.5 | 13.6 | 0.2 | -4.2 | 4.7 | 0.892 |
| *Gender* |  |  |  |  |  |  |
| Female [Reference Group] | 91.7 | 14.6 | 0.0 |  |  |  |
| Male | 88.9 | 19.4 | -2.8 | -12.6 | 7.0 | 0.470 |
| Other/unknown/prefer not to answer | 96.7 | 6.4 | 5.0 | -1.0 | 11.0 | 0.081 |
| *Race/ethnicity* |  |  |  |  |  |  |
| Non-Hispanic Black [Reference Group] | 94.2 | 12.8 | 0.0 |  |  |  |
| Hispanic/Latine | 88.2 | 17.3 | -6.1 | -13.4 | 1.3 | 0.083 |
| Multiracial | 92.3 | 12.0 | -1.9 | -7.1 | 3.3 | 0.372 |
| Other/unknown/prefer not to answer | 97.6 | 6.3 | 3.4 | -3.2 | 10.0 | 0.225 |
| *Language* |  |  |  |  |  |  |
| English [Reference Group] | 94.3 | 12.0 | 0.0 |  |  |  |
| Spanish | 86.2 | 18.2 | -8.1 | -13.1 | -3.1 | 0.011 |
| Other/unknown/prefer not to answer | 95.0 | 8.7 | 0.7 | -1.9 | 3.2 | 0.508 |
| *English proficiency* |  |  |  |  |  |  |
| Very well or well [Reference Group] | 93.9 | 12.3 | 0.0 |  |  |  |
| With difficulty | 89.0 | 14.9 | -4.9 | -9.7 | -0.2 | 0.044 |
| Unknown/prefer not to answer | 83.1 | 22.5 | -10.8 | -15.5 | -6.1 | 0.003 |
| *Pregnancy status* |  |  |  |  |  |  |
| Family [Reference Group] | 93.4 | 12.9 | 0.0 |  |  |  |
| Currently/recently pregnant | 90.5 | 15.7 | -3.2 | -7.2 | 0.9 | 0.097 |
| *Number of births* |  |  |  |  |  |  |
| None [Reference Group] | 91.6 | 15.0 | 0.0 |  |  |  |
| 1 to 2 | 94.9 | 9.8 | 3.3 | -6.5 | 13.1 | 0.405 |
| 3+ | 87.0 | 19.1 | -4.6 | -22.3 | 13.2 | 0.515 |
| Unknown/prefer not to answer/not applicable | 91.7 | 15.7 | 0.1 | -22.9 | 23.0 | 0.992 |
| *Number of preterm birth pregnancies* |  |  |  |  |  |  |
| None [Reference Group] | 93.7 | 12.8 | 0.0 |  |  |  |
| At least 1 preterm birth | 85.7 | 18.1 | -8.0 | -10.5 | -5.5 | 0.001 |
| Unknown/prefer not to answer/not applicable | 92.0 | 14.9 | -1.7 | -15.2 | 11.9 | 0.748 |
| *Pregnancy loss history* |  |  |  |  |  |  |
| None [Reference Group] | 93.0 | 13.9 | 0.0 |  |  |  |
| Prior pregnancy loss | 92.1 | 14.1 | -0.9 | -10.5 | 8.7 | 0.807 |
| Unknown/prefer not to answer/not applicable | 87.1 | 16.8 | -5.9 | -25.7 | 13.8 | 0.453 |
| *Education* |  |  |  |  |  |  |
| Less than high school degree [Reference Group] | 87.5 | 16.1 | 0.0 |  |  |  |
| High school graduate, GED, or equivalent | 91.1 | 16.2 | 3.7 | -7.8 | 15.1 | 0.424 |
| Some college, junior college, or vocational school | 97.3 | 7.7 | 9.9 | -1.3 | 21.1 | 0.071 |
| College graduate and professional or graduate school | 90.2 | 16.5 | 2.8 | -6.2 | 11.8 | 0.440 |
| Unknown/prefer not to answer | 89.8 | 14.4 | 2.4 | -1.2 | 5.9 | 0.136 |
| *Employment status* |  |  |  |  |  |  |
| Unemployed [Reference Group] | 91.5 | 14.5 | 0.0 |  |  |  |
| Full-time | 89.9 | 14.7 | -1.6 | -13.5 | 10.3 | 0.727 |
| Part-time | 98.8 | 3.0 | 7.3 | 0.5 | 14.2 | 0.041 |
| Unknown/prefer not to answer | 90.3 | 19.3 | -1.2 | -19.5 | 17.1 | 0.864 |
| *Receiving public assistance* |  |  |  |  |  |  |
| No [Reference Group] | 91.8 | 13.5 | 0.0 |  |  |  |
| Yes | 92.6 | 13.6 | 0.9 | -5.5 | 7.2 | 0.726 |
| Unknown/prefer not to answer | 88.1 | 23.3 | -3.7 | -17.2 | 9.9 | 0.492 |
| *Residence* |  |  |  |  |  |  |
| Bayview-Hunter's Point (San Francisco) [Reference Group] | 93.4 | 10.2 | 0.0 |  |  |  |
| Other San Francisco | 92.0 | 14.9 | -1.3 | -6.9 | 4.2 | 0.541 |
| East Bay | 91.3 | 12.2 | -2.0 | -8.6 | 4.5 | 0.440 |
| Other/unknown/prefer not to answer | 87.1 | 24.0 | -6.3 | -13.5 | 1.0 | 0.074 |
| *Housing status* |  |  |  |  |  |  |
| Homeless shelter [Reference Group] | 92.9 | 11.0 | 0.0 |  |  |  |
| Owens home or apartment | 99.4 | 1.7 | 6.5 | 4.8 | 8.3 | 0.001 |
| Rent home or apartment | 93.0 | 13.8 | 0.1 | -4.8 | 5.0 | 0.942 |
| Public housing | 86.8 | 18.9 | -6.0 | -20.3 | 8.3 | 0.307 |
| Unknown/Other (e.g., living with someone for free, no living place, transitional housing etc.) | 89.3 | 15.7 | -3.5 | -18.1 | 11.1 | 0.541 |
| *Social support* |  |  |  |  |  |  |
| No, not at all [Reference Group] | 87.6 | 16.4 | 0.0 |  |  |  |
| A little | 87.4 | 21.9 | -0.2 | -13.5 | 13.2 | 0.973 |
| Somewhat | 88.3 | 15.2 | 0.7 | -11.6 | 13.0 | 0.884 |
| Yes, definitely | 94.8 | 11.7 | 7.2 | -6.7 | 21.1 | 0.224 |
| Unknown/prefer not to answer | 94.4 | 9.6 | 6.8 | -10.1 | 23.7 | 0.325 |
| *Medical Insurance* |  |  |  |  |  |  |
| Public insurance (e.g., e.g., Medi-Cal, Medicaid, etc.) [Reference Group] | 95.1 | 10.2 | 0.0 |  |  |  |
| Private or employer-provided insurance | 89.7 | 14.1 | -5.4 | -17.6 | 6.7 | 0.284 |
| No insurance | 77.1 | 19.7 | -18.0 | -29.0 | -6.9 | 0.011 |
| Unknown/prefer not to answer | 82.5 | 23.9 | -12.6 | -24.1 | -1.2 | 0.038 |
| *Food insecurity (Worried food would run out)* |  |  |  |  |  |  |
| Often true [Reference Group] | 90.1 | 13.7 | 0.0 |  |  |  |
| Sometimes true | 93.8 | 10.0 | 3.7 | -5.1 | 12.5 | 0.308 |
| Never true | 95.4 | 10.1 | 5.3 | 0.3 | 10.3 | 0.042 |
| Unknown/prefer not to answer | 78.6 | 23.8 | -11.5 | -16.3 | -6.8 | 0.003 |
| *Food insecurity (Insufficient funds)* |  |  |  |  |  |  |
| Often true [Reference Group] | 90.5 | 13.5 | 0.0 |  |  |  |
| Sometimes true | 91.2 | 12.7 | 0.7 | -17.1 | 18.5 | 0.919 |
| Never true | 94.8 | 12.1 | 4.3 | -3.0 | 11.6 | 0.179 |
| Unknown/prefer not to answer | 86.6 | 21.2 | -3.9 | -15.9 | 8.1 | 0.419 |
| *Relationship status* |  |  |  |  |  |  |
| Married/partnered, living together [Reference Group] | 90.3 | 13.0 | 0.0 |  |  |  |
| Married/partnered, not living together | 92.6 | 16.9 | 2.3 | -17.2 | 21.7 | 0.763 |
| Single | 95.1 | 10.0 | 4.8 | -3.3 | 13.0 | 0.175 |
| Other/unknown/prefer not to answer | 82.7 | 24.6 | -7.6 | -24.8 | 9.7 | 0.290 |
| *Everyday discrimination* |  |  |  |  |  |  |
| Never [Reference Group] | 93.3 | 13.9 | 0.0 |  |  |  |
| Rarely | 93.4 | 11.2 | 0.1 | -6.7 | 6.9 | 0.968 |
| Sometimes | 90.1 | 17.5 | -3.2 | -7.2 | 0.7 | 0.086 |
| Often | 92.2 | 11.5 | -1.1 | -2.3 | 0.1 | 0.068 |
| *Discrimination during prenatal care encounters* |  |  |  |  |  |  |
| Never [Reference Group] | 96.8 | 7.4 | 0.0 |  |  |  |
| Rarely | 90.2 | 14.7 | -6.6 | -8.4 | -4.8 | 0.000 |
| Sometimes | 85.4 | 19.2 | -11.4 | -20.0 | -2.9 | 0.021 |
| Often | 95.2 | 11.7 | -1.6 | -4.7 | 1.6 | 0.237 |

Abbreviations: 95% CI: 95% confidence interval; SD: standard deviation

In the final multivariate model, acceptability was significantly associated with language, medical insurance, and experience of discrimination during prenatal care encounters (Table 6). Participants who spoke Spanish found PV less acceptable than those who spoke English, scoring, on average, 7.9 points lower (95% CI −15.6, −0.2). On average, participants without medical insurance scored, on average, 16.3 points (95%CI −28.0, −4.7) lower than those who had public insurance. Participants who experienced discrimination during the usual prenatal care encounters found PV less acceptable than those who did not experience any discrimination during care encounters, scoring on average 10.3 points lower (95% CI: −15.5, −5.0).

**Table 6.** Multivariate mixed-effect linear regression model of select predictors on the Acceptability score.

| Predictor variables |  |  |  |  |
| --- | --- | --- | --- | --- |
|  | Coeff. | [95% CI] |  | P-value |
| Language |  |  |  |  |
| English [Reference Group] | 0.0 |  |  |  |
| Spanish | -7.9 | -15.6 | -0.2 | 0.044 |
| Other/unknown/prefer not to answer | 2.4 | -7.5 | 12.3 | 0.629 |
| English proficiency |  |  |  |  |
| Very well or well [Reference Group] | 0.0 |  |  |  |
| With difficulty | 3.5 | -5.4 | 12.3 | 0.441 |
| Unknown/prefer not to answer | -0.8 | -10.5 | 8.9 | 0.874 |
| *Number of preterm birth pregnancies* |  |  |  |  |
| None [Reference Group] | 0.0 |  |  |  |
| At least 1 preterm birth | -4.3 | -10.2 | 1.6 | 0.157 |
| Unknown/prefer not to answer/not applicable | 8.2 | -0.4 | 16.7 | 0.061 |
| *Employment status* |  |  |  |  |
| Unemployed [Reference Group] | 0.0 |  |  |  |
| Full-time | -0.6 | -8.8 | 7.6 | 0.888 |
| Part-time | 4.2 | -3.4 | 11.8 | 0.282 |
| Unknown/prefer not to answer | 0.0 | -8.0 | 8.0 | 0.993 |
| *Housing status* |  |  |  |  |
| Homeless shelter [Reference Group] | 0.0 |  |  |  |
| Owns home or apartment | 7.2 | -3.5 | 17.8 | 0.189 |
| Rent home or apartment | 4.4 | -3.7 | 12.5 | 0.288 |
| Public housing | -1.2 | -10.2 | 7.9 | 0.802 |
| Unknown/Other (e.g., living with someone for free, no living place, transitional housing, etc.) | 3.8 | -4.6 | 12.3 | 0.378 |
| *Medical Insurance* |  |  |  |  |
| Public insurance (e.g., Medi-Cal, Medicaid, etc.) [Reference Group] | 0.0 |  |  |  |
| Private or employer-provided insurance | -3.0 | -10.4 | 4.4 | 0.425 |
| No insurance | -16.3 | -28.0 | -4.7 | 0.006 |
| Unknown/prefer not to answer | -6.6 | -14.1 | 1.0 | 0.09 |
| *Food insecurity (Worried food would run out)* |  |  |  |  |
| Often true [Reference Group] | 0.0 |  |  |  |
| Sometimes true | 4.1 | -3.5 | 11.8 | 0.288 |
| Never true | 1.8 | -5.6 | 9.1 | 0.640 |
| Unknown/prefer not to answer | -7.8 | -16.4 | 0.8 | 0.077 |
| *Discrimination during prenatal care encounters* |  |  |  |  |
| Never [Reference Group] | 0.0 |  |  |  |
| Rarely | -7.6 | -13.8 | -1.4 | 0.017 |
| Sometimes | -10.3 | -15.5 | -5.0 | 0.000 |
| Often | -5.4 | -14.4 | 3.5 | 0.234 |

Abbreviations: 95% CI: 95% confidence interval; SD: standard deviation

Participants’ responses from the in-depth interviews supported the quantitative data on acceptability. We summarized the data on the acceptability of PV using six deductive domains based on the TFA.^14^ The definitions of the TFA domains and additional excerpts of the outlined domains are shown in Figure 1.

**Figure 1.**
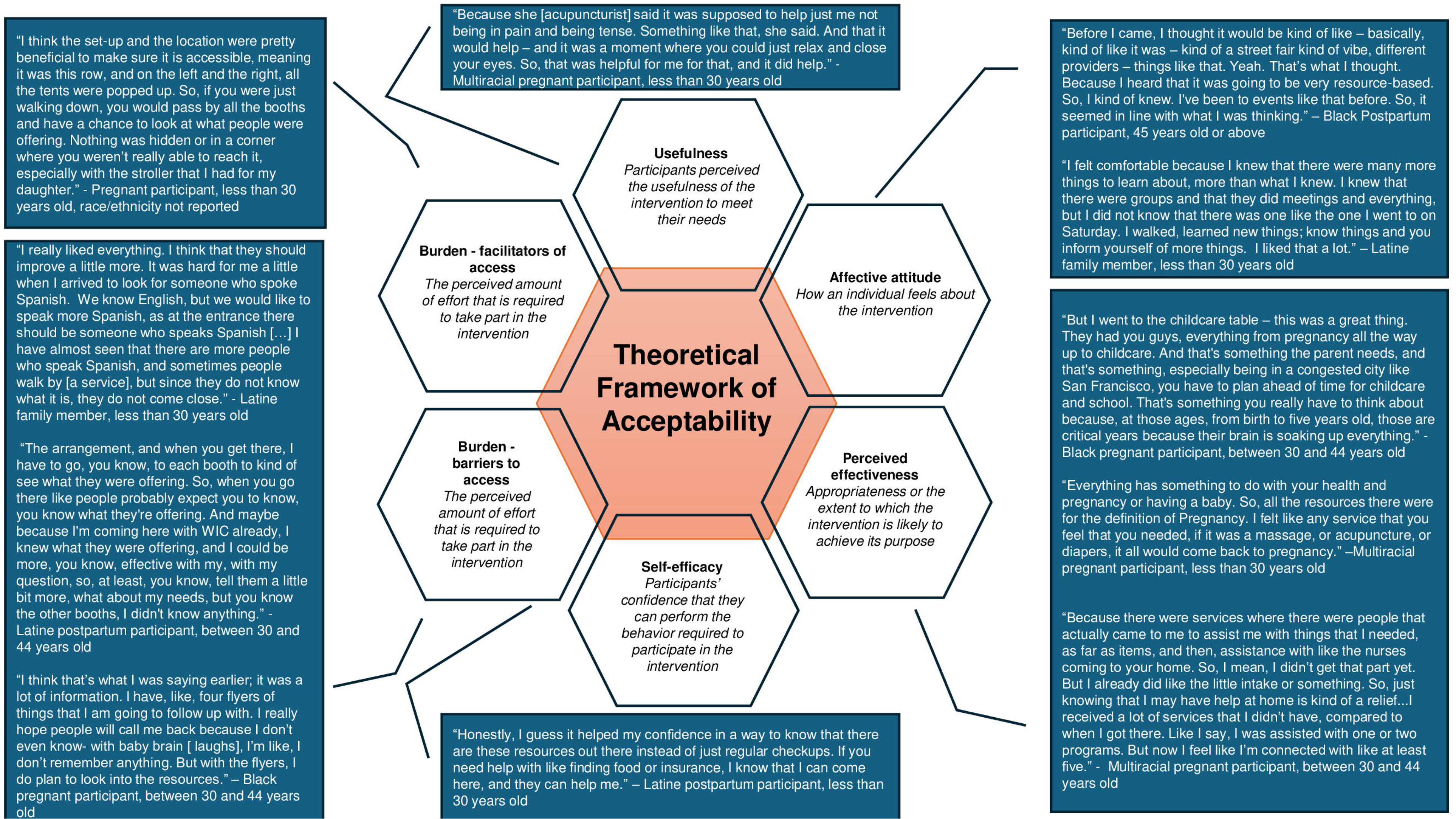
Theoretical Framework of Acceptability (TFA) domains and corresponding quotes. Designed by Jorrín Abellán, I.M. Hopscotch Building: A Model for the Generation of Qualitative Research Designs (2016).^15^. This figure displays the domains of the Theoretical Framework of Acceptability, paired with illustrative quotes that capture participants’ perspectives on each domain.

#### Anticipated affective attitude

When asked about their expectations of PV prior to attending an event, some participants noted they expected a health fair-type venue to receive resources and information about pregnancy. When reflecting on their experience at PV, participants indicated that their expectations were met, noting the inviting and uplifting atmosphere and the abundance of resources—including mental health and wellness services like massages and acupuncture, and prenatal offerings such as breastfeeding education. Many also noted that they gained valuable awareness and knowledge about various services designed to improve their pregnancy experience.

> *“[I expected PV to be] kind of similar to what it is. So, definitely, organizations and businesses that have resources for pregnant people. I didn’t expect there to be that many giveaways for pregnant people or services in terms of massages. So, more just like resources, knowing like, ‘Hey, this is where you can get XYZ. This is available to you here in San Francisco.’ And the education part, I kind of did expect, like the breast-feeding education.”* —Pregnant participant, less than 30 years old, race/ethnicity not reported

A few participants, however, did not have any expectations of the event as they were unfamiliar with community-centered events where individuals could learn about and receive resources. For example, one family member noted:

> *“Well, you know, I didn’t have any expectations. I never heard of anything like that, never been to a community gathering of that sort. So, I didn’t have a whole lot of expectations. But, as I read the flyer and read the different organizations that were going to be there, I kind of felt good about attending and felt like it was worthwhile.”* —Black family member, 45 years old or above

#### Experienced affective attitude

In general, participants described the PV atmosphere as positive, vibrant, community-centered, down-to-earth, and healing. When asked how they felt upon arriving at PV, participants expressed feeling welcomed, excited, and filled with anticipation. For instance, one Black postpartum participant had this to say:

> *“I was happy. I was like, ‘Wow. Look at all of this.’[…] because I had my son there – he’s seven-and-a-half months. And I was like, “Look!” He really loved the color of the tents and the way the tents were kind of blowing in the wind. He loved the live music. It was just like, “Wow. Okay. Yes!” And it was like it was our first time being out in community ever. And so, I was pleased that it was such a warm, welcoming, fun environment for us to walk into. You know? You walk in and you see like the bags of groceries that, you know, were being handed out and things like that.”* —Black postpartum participant, 45 years old or above

Participants also expressed their delight and appreciation for the new services and resources that were added at each event, which was unexpected.

> *“I was excited to see what was new - because you guys don’t always have the same things; sometimes what you all give away and what you all do for us is unexpected, especially for the resources. I didn’t think the CalWORKS individuals were going to be there for the moms that don’t have any type of income.”* —Multiracial pregnant participant, age not reported

Some participants recalled that their initial impressions of PV were that it felt like a community-centered, curated space where Black individuals could come together to enjoy each other’s company and learn about pregnancy-related services and resources. They shared that being surrounded by other Black individuals or expectant mothers made them feel at home—free to express themselves and engage in activities without fear of judgment.

> *“I felt kind of like at home. Because I wasn’t like the only one pregnant, or there were mothers that were there that were very comfortable breastfeeding and stuff. So, seeing that was like, oh, this is a place for us to just be open and free without no judgment.”* — Multiracial Pregnant participant, 30-44 years old

> *“It was about seeing Black individuals coming together without fussing or arguing. We were there, we enjoyed each other, we laughed. That makes you feel good when Black individuals come together and everybody enjoys themselves […] “I looked around. I’m like, ‘Wow, this is neat! This could help a lot of young girls.’ You know, teach them. That’s what we need to do is teach them the right way.”* —Black family member participant, 45 years old or above

The community-centeredness of PV was especially relevant in the context of the social isolation individuals felt during the COVID-19 pandemic, as noted by this participant:

> *“I was pregnant from 2020 all the way through 2021. That was like the height of when everything was like shut down and isolated. We were having fires. I couldn’t go outside to walk even, hardly. It was extremely isolating. And so, one of the benefits that I don’t think that you all are necessarily picking up on is that, for individuals who have been isolated because you’re trying to follow COVID protocols and all of that, it was healing to just be out in the fresh air, in the sunshine, with other mothers. Like that, in and of itself, was the healing thing for me.”* —Black postpartum participant, 45 years old or above

Participants reflected that attending PV had made them more aware of shared interests with others in their community, which for some, had given rise to a desire to become more actively involved in their community:

> *“Going to the pop-up just opened my eyes to there’s so many individuals out there that I have common ground with. And we can all work on ourselves together. We could help each other. Some individuals don’t know where to start. So, that’s where I’m at. It kind of inspired me to definitely see what I could do for my community. But first, yeah, I felt very good.”* —Latine postpartum participant, between 30 and 44 years old

Most participants said their expectations were met, with some acknowledging that their expectations were exceeded. Participants cited the plethora of resources and services, including resources that catered to physical, psychological, and spiritual enrichment, and the feeling of being valued and acknowledged, as reasons for their expectations being met.

> *“[My expectations] were met. I received a lot of information from different groups on childcare, doula services, even when the baby starts growing her first teeth, even a dentist that’s nearby my house. And chiropractic services for pregnant women – I didn’t even know you can get realigned as a pregnant woman.”* —Black pregnant participant between 30 and 44 years old

Others noted that their expectations were met because they felt acknowledged and valued at the PV, as one pregnant participant shared: *“I really felt acknowledged and seen, and individuals were really trying to take care of me because they saw that I was pregnant.” —*Pregnant participant, less than 30 years old, race/ethnicity not reported

However, a few participants reported that their expectations were not met due to having inadequate guidance regarding the resources available to them.

> *“Yeah, so once again I have, you know, totally different expectations., So when I went there, and I see just a bulletin and nobody, nobody is, like, welcoming you, you know. But, if someone doesn’t know anything, like where do you go? I just had to ask around, and they told me, like, ‘Okay, you can get some information here in that first booth.’ But I didn’t really get that information well.”* —Latine postpartum participant, between 30 and 44 years old

#### Self-efficacy

Overall, participants were very confident accessing services at PV, as well as outside of PV, after attending. Participants attributed their increased confidence to PV’s strong referral system, learning about new services, and receiving adequate information from service providers:

> *“I feel like it was a linkage sort of situation. So that, if I needed something more extensive, then I would have the information to know like where to go, who to talk to – all of that – maybe even set up an appointment. So, when I did need to go and access that deeper level of services, I could go out into the community, and then do that.”* —Black postpartum participant, 45 years or above

However, a few participants shared that PV did not increase their confidence in accessing new services, primarily because they were unaware of the full range of services offered or only received information from organizations where they were already enrolled, limiting their exposure to new services:

> *“I don’t think so, I mean, the only, the only things that I saw that was convenient for me was the programs that I already know. But anything else that I don’t know, I don’t know. The Homeless Prenatal, like I’ve been a customer before. And then for, for WIC, you know, I’m a client already. So, anything else, I have no idea.”* —Latine pregnant person between 30 and 44 years old

#### Appropriateness

Participants generally found the services appropriate based on their expectations. Some participants perceived services to be appropriate because they were aimed at meeting the needs of pregnant individuals and their families:

> *“I feel like they’re appropriate because the need is so great right now. A lot of individuals are on hard times. A lot of individuals are struggling. And, with these organizations being implemented the way they have to help families, it just kind of lightens the load. It gives me just that great feeling of knowing – even though I’m on hard times right now – that I don’t have to worry because I can get the help that I need until I’m in a better place and can do better for myself. So, I feel like it’s totally appropriate.”* —Black family member, 45 years old or above

Participants felt the services were appropriate because they provided education about pregnancy, including filling gaps in breastfeeding education:

> *“Even though I had already been through breastfeeding when I first started with my first child, I realized it is really something that women need support on. Even though it is natural, it doesn’t come naturally to a lot of people. And just knowing that there was this booth that really educated individuals on the latch, and how breast milk is established, and that it might take a while.”* —Pregnant participant, less than 30 years old, race/ethnicity not reported

A few participants, however, felt that some services were inappropriate, citing a perceived lack of pregnancy-related services. As one pregnant participant explained:

> *“I went for information for pregnancy, and when I went there, I didn’t find anything kind of related, at least, you know, with my eye at the beginning. And then I guess it was a couple of books for that, but I didn’t see anybody or was expecting to see more people, you know, kind of with a similar concern that I have, and I see nobody.”* —Latine pregnant participant between 30 and 44 years old

#### Perceived effectiveness

Participants generally perceived PV to be effective, noting its uplifting environment and focus on centering Black women.

> *“I think they did a good job of providing that really positive atmosphere. Especially I feel like it was centering around women of color and Black women, and sometimes we will often feel like society look down, like “oh you are pregnant and [inaudible].” I think they did a good job of making it feel like a positive thing you’re pregnant.”* —Black pregnant participant, between 30 and 44 years old

Other participants attributed the effectiveness of PV to its ability to meet the needs of the community, such as outreaching to community and delivering resources in an accessible way:

> *“I like that you guys like to be part of the community, and you like to see what is going on in the community. It is really hard right now, especially to be homeless, and not have a lot of resources - that’s one thing I like - for you guys to go and give back to the community and say ‘Hey, we have resources. We want you guys to come out and have a good time as well.’ And on top of that, it also helps with depression.”* —Multiracial pregnant participant, age not reported

#### Usefulness

Participants generally found the services and resources provided at PV to be useful. Some participants cited pregnancy and postpartum-related education, such as newborn hygiene and nutrition, as well as mental health support, as key reasons for its perceived usefulness.

> *“Well, at the one table that was in the middle, I overheard them talking about how to give baby a bath, how to do this with your baby, do that with your baby. That was so cool because I saw individuals with babies and stuff.] I was kind of listening a little bit. They were sitting there talking about their kids, what their kids do, what their kids eat, what they didn’t eat, what’s good for them and what’s not good for them. And that’s what they need. They need to know more about baby nutrition.”* —Black family member, 45 years old or above

Several participants found PV useful because they offered educational resources for children and children-related activities at the events.

> *“I saw how they were with the kids. The little kids were up there drawing, doing all this stuff. You know, they were really, I think they were really supportive, helping all the little kids up there doing the thing.”* —Black family member, 45 years old or above

### Service recommendations

In the in-depth interviews, participants shared several recommendations to improve the PV model. These included adding or improving specific services, such as provider follow-up with interested individuals; provision of more food, particularly for pregnant individuals; expanded pregnancy education sessions; more support and education around mental health, including postpartum depression and managing the stress of newborn care; and additional child-focused activities and resources, particularly those that allow parents and children to engage together.

> *“I think they did have a booth for, like, mental health support. But definitely more education around health and depression, and the fourth trimester, and post-partum essentials that are really helpful for a mother for recovery, and then also for a baby. Just that it’s not just about the pregnancy or when you give birth, but then the first few weeks or the month after that might be really intense on women.”* —Black pregnant participant, less than 30 years old

> *“If you guys have like some sort of guided mommy and me activity like with music and stuff like that, where you could just kind of come over with your baby and be led in some like movement activities and stuff. That would be fun.”* —Black postpartum participant, 45 years old and above

Others shared infrastructural recommendations, such as providing more comfortable seating areas, engaging navigators to guide visitors throughout the Village and inform them about relevant services—particularly in their preferred language—and increasing outreach to ensure broader community awareness of available services.

### Sensitivity analyses

In sensitivity analyses in which missing and duplicate responses were excluded, we identified no differences in standardized acceptability or accessibility scores (see Supplementary Table 3). The acceptability score was 90.8 (SD 15.2) overall (*N*=85), 90.5 (SD 15.9) for pregnant and postpartum participants (N=45), and 91.2 (SD 14.5) for family members (N=40). The accessibility score was 74.5 (SD 22.6) for the main sample (N = 87), 71.0 (SD = 24.2) for pregnant and postpartum participants (N=46), and 78.5 (SD 20.2) for family members (N=41).

## DISCUSSION

Participants’ perceptions of PV’s accessibility and acceptability were high, suggesting the Pregnancy Village model is accessible and well-accepted by the community. Accessibility and acceptability were both patterned by language proficiency, socioeconomic status, and discrimination experiences within prenatal care. Qualitative findings complemented and expanded the quantitative results, highlighting specific factors that influenced participants’ assessment of accessibility and acceptability. These findings support the model as accessible and acceptable, offering a pathway to improve the experiences of Black and other minoritized pregnant and postpartum individuals and their families, and offer tangible strategies for improvement.

Quantitative findings on the accessibility of the PV suggest some room for improvement. One reason for this is that although PV was focused on SF residents, some participants came from other parts of the Bay Area. Thus, the time to reach events was longer for these individuals, who aren’t the target population for the program. The main barriers to accessibility cited in the qualitative interviews were, however, related to the burden of participation, as in the effort required by participants to participate in and access PV services once at the location. Although the resources provided were viewed as valuable and useful, some participants noted difficulty navigating and interacting with the resources, which suggests a need to optimize service linkage and delivery, even in community care settings such as PV. Some participants, particularly Spanish-speaking participants, had lower perceptions of the events’ accessibility due to the language barrier, making it more difficult for participants to obtain the services they needed. This is consistent with research indicating that a language barrier is an obstacle among migrant and diverse women who seek prenatal care.^16,17^

The high acceptability of the PV was reflected in participants’ positive feelings about the PV’s vibrant, welcoming, and healing atmosphere. Literature on the impact of the built environment on emotions suggests that the human experience is shaped by how space affects mood, comfort levels, and engagement with their environment, as well as by architectural elements (e.g., openness and connectivity of spaces).^18–20^ PV events are held outdoors in spaces with an open layout, featuring brightly colored tents and seating areas, along with a variety of activities and services. Restorative elements such as natural daylight, openness, an aesthetically pleasing layout, and exposure to nature are known to enhance perceived restorativeness, likely contributing to participants’ strong acceptance of PV.^20^ A welcoming environment is one where individuals feel both represented and acknowledged, treated with respect and dignity, and where all cultural identities are valued (New York State Education Department, *Culturally Responsive-Sustaining Education*, https://www.nysed.gov/sites/default/files/programs/crs/culturally-responsive-sustaining-education-framework.pdf). One key reason participants attribute high acceptability to the welcoming environment may be that it fosters a sense of hope. By providing opportunities for people to meet new individuals, learn new things, and improve their health and wellness,^21^ PV creates an atmosphere of positive change. Engaging in such activities has the potential to improve one’s affect and overall health and wellness, suggesting that fostering a welcoming environment is crucial for improving people’s affective attitude. In turn, this may increase the perceived acceptability of a community sociomedical intervention like PV.^21–23^

Participants also reported that the community-centeredness of PV contributed to their acceptance of the model, particularly in light of the shutdowns and social distancing measures implemented during the COVID-19 pandemic, which led to increased social isolation. PV visitors likely experienced social support while observing and practicing health-promoting behaviors such as participating in food demonstrations, dance classes, yoga classes, and sharing circles, all of which are regularly provided, thereby influencing their perceived acceptance of PV.^24–26^ Research has demonstrated that a strong sense of community is positively associated with problem-focused coping practices that strive to address underlying issues, such as lack of access to resources, while a lack of a sense of community has been associated with reduced use of these constructive coping behaviors.^24,33,34^ Additionally, a sense of community has been demonstrated to be positively associated with an increased ability to perform health-promoting behaviors, which may contribute to participants’ acceptance of PV.^29^

Participants attributed their acceptance of PV to its ability to increase their confidence in accessing and utilizing available resources both within and outside the PV events, implying that the intervention promoted self-efficacy. Learning about resources and having enough information about resources increased participants’ perceived self-efficacy. This finding is congruent with a meta-analysis that found public health interventions—aimed at disadvantaged populations—that employed community engagement strategies significantly increased participants’ self-efficacy.^35^ This may be explained in part by the health literacy-focused activities provided at events like PV, which have been linked to increased perceived self-efficacy in several studies.^36–38^ However, it is also possible that individuals who attended PV events are more likely to be self-efficacious in general, given previous research^30,39^ demonstrating that individuals with higher self-efficacy are more likely to engage in health-promoting behaviors, such as participating in interventions like PV events;^31,40^ this may be another reason why participants accept the model.

Our quantitative findings also revealed that participants who experienced discrimination during prenatal care encounters scored significantly lower on acceptability than those who did not. One possible explanation for this significant association is that participants’ earlier experiences with discrimination while seeking care or prior unfavorable experiences with organizations that provide services at PV may have influenced their perceived acceptability of the events. This is supported by previous studies. For example, one qualitative study found that earlier experiences of discrimination were a reason why individuals rejected social care interventions.^32^ As a result, providing culturally sensitive care is critical to dispelling misapprehensions about perinatal care.

Participants suggested continued improvements to PV. First, post-event follow-up with participants who are interested in learning about or receiving services is important to make services more accessible. Establishing a structured method for consistent provider follow-up is critical to ensure PV remains a trusted resource within the community. Second, PV should have navigators who can advise and guide visitors to services that may be of interest to them. Third, the presence of translators and/or Spanish-speaking PV providers was an important suggestion, as language-concordant care has been linked to enhanced communication, higher satisfaction in care, engagement in care, and improved health outcomes.^41^ Fourth, PV should provide lunch or other food provisions to ensure that all visitors are fed. A study has found that the provision of food can address meeting community food needs and food access (UW Center for Cooperatives, *Consumer Cooperatives*, https://uwcc.wisc.edu/resources/consumer-cooperatives/), foster community engagement,^42^ and encourage engagement in health-promoting behaviors.^43^ Fifth, participants advocated for greater perinatal education. Prenatal education has been shown in studies to minimize childbirth anxiety^44–46^ and depression, and boost childbirth self-efficacy.^46–49^ Sixth, participants suggested that PV increase its community outreach efforts, particularly to its priority population of Black birthing and postpartum individuals and their families. Other community-based ‘fair’ initiatives aimed at underserved populations have identified the need for investments in advocacy and targeted outreach to optimize the care received.^50^ These improvement suggestions also emerged through the community feedback mechanisms built into the PV and were later incorporated into the events, highlighting the importance of building such mechanisms for feedback and community co-design.

This study has some limitations. First, because the evaluation was in the context of real-world implementation and the model’s dynamic co-creation approach allowed it to adapt to the expressed needs of the participants, the model being evaluated was inconsistent over time. Given that the PV events were designed to respond to community needs, the factors cited as barriers to access or reasons for participants’ perceived low acceptance of the model became less prominent as the evaluation continued. Second, our purposive and convenience sample limits generalizability to other populations. Third, completing the survey onsite by some participants may have introduced social desirability bias. To mitigate this, surveys were self-administered and anonymous, with assurances that responses would remain confidential and have no impact on participants’ involvement in PV events. In addition, given that participants could respond multiple times if they attended multiple events, the data may overrepresent the experiences of those who had positive experiences and returned to subsequent events. Sensitivity analysis, however, suggests this is unlikely since the results did not change significantly when restricted to only the responses from unique individuals.

The study has several strengths. First, is the use of a mixed methods design: the in-depth interviews enabled us to gain a more nuanced understanding of participant perceptions across acceptability domains than was feasible with the quantitative assessment alone. Second, is the use of the Theoretical Framework of Acceptability as an orienting framework. We identified two additional acceptability domains in our study (i.e., perceived usefulness and appropriateness), which are not captured in the TFA domains. We, therefore, suggest modifications to the TFA to include these domains that may be useful for other researchers evaluating the acceptability of community sociomedical interventions. Further, by involving key partners in the development of data collection tools, including community members and the implementation team, we were able to identify questions that were relevant to both participants and those designing the events.

Reducing disparities in maternal and neonatal health outcomes requires improving access to a wide range of perinatal support and care needs beyond those that are traditionally provided within the healthcare setting and doing so in a way that is culturally appropriate and acceptable. Our study demonstrates that providing one-stop shop access to diverse services within a community-centered and uplifting setting is an acceptable approach to meeting community needs, but supporting and sustaining such programs will require policies that take a holistic, integrated, and community-centered approach to improving health.^51,52^ One policy strategy is to incentivize community-institutional collaborations that encourage integration and shift away from the current standard of services being delivered independently by different organizations with little coordination.^51,53^ One approach may be through novel types of funding; for example, California’s CalAIM Population Health Management program requires managed Medicaid health plans to conduct regular community needs assessments within local health jurisdictions and to invest in community health improvement projects that address upstream drivers of health through integration of healthcare with public health and social services and build trust with and meaningfully engage members (Department of Health Care Services, *CalAIM: Population Health Management (PHM) Policy Guide*, 2024). Longer-term meaningful collaborations may also entail redesigning health and social systems to overcome structural hurdles and simplifying access to equitable care, which is the overarching goal of the “Pregnancy Village” vision.

Our study also highlights the importance of critical implementation practices for programs focused on reducing health inequities. To ensure programs continuously meet participants’ needs, it is critical to incorporate processes for inviting continuous feedback from participants and iterating in response. Many of the improvement needs identified through our survey and interview data also emerged in the feedback mechanisms built into our events, such as feedback boards and other activities,^13^ which allowed for responsive changes to be made in real time. Our findings that participants needed support with navigation even within our setting of service co-location, had challenges with access due to lack of transportation, and felt that food provision at events was critical, highlight how individuals of color and those living on low incomes are impacted by structural inequities; and deep reflection and an intentional anti-racism lens is required to ensure new programs are designed and implemented in a way that tackles rather than propagates these inequities.

Intervention accessibility and acceptability are critical for participant engagement and adherence, as well as the sustainability of an intervention. The experiences and views of pregnant or postpartum individuals and family members who participated in the PV events were utilized to assess the accessibility and acceptability of this cross-sector, community-engaged care model. As the first iteration of the Pregnancy Village model, the SF Family & Pregnancy Pop-Up Village was found to be accessible and acceptable for Black and other minoritized pregnant individuals and their families living in the San Francisco Bay Area, California. Continued community-institutional co-creation is a priority to ensure ongoing integration of community priorities. Future work will robustly assess the impact of the Pregnancy Village model on care access, experience, and mental well-being, and eventually, on health outcomes. Ongoing process and planned impact evaluation of this model will inform the transportability of this community-responsive model for service provision and support in other contexts.

## METHODS

### Setting

San Francisco’s Bayview district was chosen as the PV site as it is home to 19% of Medicaid-insured and 36% Black-identifying birthing SF residents (California Department of Public Health, *Vital Records Business Intelligence System (VRBIS)*, 2022)). Most birthing individuals in the Bayview are Medicaid-insured (61%), and 93% identify as members of a racial or ethnic minority group (SFDPH MCAH, *SFDPH Tableau Data*, 2021). Furthermore, Bayview residents have significantly lower rates of timely prenatal care (inequities in access) and higher rates of preterm birth (inequities in outcomes) than residents of other neighborhoods, even among Medicaid-insured individuals only (California Department of Public Health, *Vital Records Business Intelligence System (VRBIS)*, 2022)).

The Bayview combines industrial, commercial, and residential zoning areas, but industrial shifts have caused infrastructure disinvestment over time. Car traffic is higher than foot due to Bayview’s hilly terrain, few street trees, and accessible parks. PV events were initially centrally located near a major thoroughfare with public transportation and community institutions. Proximity to trusted community institutions was intentional to encourage community trust and engage these organizations. Vibrant colors, shade, ground treatment, and diverse seating were incorporated into the event design to transform the area into an active and welcoming space that represented PV values.

### Intervention

The Pregnancy Village model of cross-sector collaboration aims to address perinatal inequities by providing a one-stop shop for comprehensive care and support in a dignified and empowering environment tailored to Black pregnant individuals and their families. Monthly events bring together various partners, including city government, healthcare, and community-based organizations. The range of services provided includes healthcare (e.g., Medicaid enrollment and consultations with medical professionals), as well as holistic wellness services (e.g., dance and cooking demonstrations, acupuncture, massage, and sharing circles). The model is rooted in anti-racism and person-centered care principles, prioritizes sustainable community-institutional partnerships, and includes a real-time community feedback mechanism to ensure continuous model improvement (Figure 2). More information about its implementation can be found elsewhere.^13^

**Figure 2.**
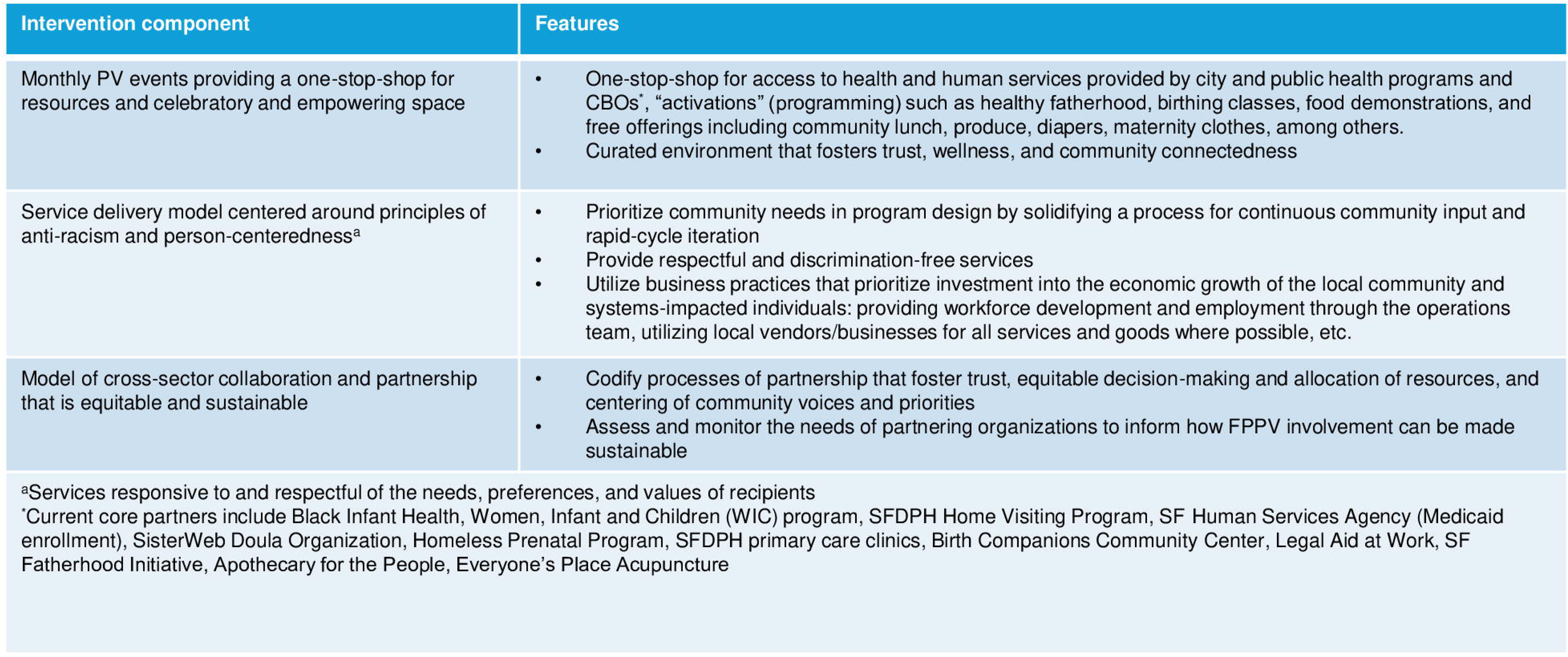
Pregnancy Village Model Components and Features. This figure illustrates the core components of the Pregnancy Village model, designed to provide a one-stop shop of cross-sector services in a safe, healing, and uplifting environment for pregnant individuals and their families.

### Study design

We employed a convergent mixed-methods, community-engaged process evaluation involving triangulation of qualitative and quantitative data to assess: 1) the feasibility and fidelity of the PV model; 2) accessibility and acceptability of PV among participants; 3) factors affecting the sustainability of provider participation; and 4) the preliminary impact of PV, such as perceptions of person-centeredness, comfort, and trust. This paper focuses on the findings related to accessibility and acceptability.

### Sample

We recruited a convenience sample of pregnant and postpartum individuals and family members from the first nine PV events between July 2021 and June 2022. This period was selected because it signifies the early phase of PV, when both participants and providers were adapting to novel processes within a co-led community-institutional model, and several adaptations to the model were being made in response to emerging needs and feedback. Capturing and examining these early dynamics was therefore critical to understanding how the model functioned in its first iteration, how participants perceived the care environment, and how these early experiences might shape longer-term engagement and outcomes. Although the Pregnancy Village model and resulting SF Family & Pregnancy Pop-Up Village events focused on improving the perinatal experience and outcomes of Black individuals and were designed accordingly, the organizers believed that the events would also be appealing and beneficial to other pregnant individuals and families facing barriers to care, such as other minoritized groups and/or those living on low incomes. We, therefore, targeted Black and other minoritized pregnant or postpartum individuals and their families who were receiving services at PV for the evaluation. Inclusion criteria were: (1) at least 15 years old if pregnant or postpartum, or 18 years old if family; (2) participation in at least one PV event; and (3) able to speak English or Spanish. We aimed to recruit 120 individuals in total, targeting a feasibility number of 10-15 from each event. Eligible participants who attended multiple PV events were allowed to participate multiple times. We enrolled 120 individuals for surveys and purposively selected a subset of participants (n=18) for semi-structured in-depth interviews, maintaining a 70-30 ratio between pregnant and postpartum individuals (n=13) and family members (n=5). We employed a purposive sampling strategy informed by the principle of maximum variation, with the goal of capturing a diverse range of experiences by considering factors such as race/ethnicity, pregnancy status, language, and age.^54^ Interviews were conducted until thematic saturation was achieved, as no new themes emerged in the final interviews.

### Recruitment and data collection

#### Quantitative

During registration at PV events, potentially eligible participants were invited to learn more about a study evaluating their experiences. Study team members provided information about the study, screened interested individuals for eligibility, and invited those who were eligible to participate. After obtaining verbal informed consent, participants were invited to complete a survey about their experiences onsite using a study tablet or later using their own devices via a QR code; most opted to do it on-site. The survey assessed various domains of the PV experience. Questions were adopted or adapted from validated scales or developed by the study team with input from a community advisory board (CAB), informed by cognitive interviews with three pregnant and postpartum individuals from the Bayview and the greater San Francisco Bay Area. Participants received a $20 gift card as compensation for completing the survey.

#### Qualitative

Three to four survey participants from each PV event were invited to take part in in-depth interviews. The interviews were scheduled at times that suited the participants. The semi-structured interview guide was informed by the Theoretical Framework of Acceptability (TFA) and included additional questions on accessibility, person-centered care, community engagement, and trust in the healthcare system. Interviews lasted 30 to 60 minutes and were conducted via Zoom in either English or Spanish; participants could join through the Zoom app or by phone, ensuring access regardless of internet or device limitations. Interviews took place within four weeks of attending a PV event and were conducted by researchers with qualitative training (OJO and KV). In-depth interview participants were compensated an additional $20. In-depth interviews were recorded with participant permission and transcribed for analysis by a third-party transcriptionist. Transcripts were checked for accuracy and clarity by members of the study team (KV and JV). Field notes were taken during the interviews, and a templated summary outline was created afterward, facilitating rapid analysis for model iteration.^13^

### Quantitative Measures

#### Accessibility

Participants’ perceptions of PV accessibility were assessed through an accessibility scale composed of three questions based on investigators’ prior experience conducting research with this target population (Supplementary Table 4 1): (1) “How long did it take you to get to PV from where you typically stay?” (2) “How do you feel about the time it took you to get to PV?” (3) “Compared to how you access your usual sources of care and resources, would you say the services at Pop-Up Village are…?” The responses to the three items were summed and then standardized (range 0-100), with higher scores indicating higher accessibility. Missing data (3.4%) were imputed as the mean of other items in the measure. Internal consistency reliability for accessibility within this sample was Cronbach’s α = 0.53

#### Acceptability

Acceptability was assessed using a 7-item scale adapted from the acceptability of health apps among adolescents (AHAA) scale for the acceptability of health interventions.^14,55^ We assessed the following sub-domains of TFA: (1) affective attitude (i.e., how an individual feels about the intervention); (2) burden (i.e., the perceived amount of effort that is required to take part in the intervention; (3) intervention coherence (i.e., the extent to which the participant understands the intervention and how it works); (4) opportunity costs (i.e., the extent to which benefits, profits, or values must be given up to engage in the intervention); (5) perceived effectiveness (i.e., appropriateness or the extent to which the intervention is likely to achieve its purpose); and (6) self-efficacy (i.e., participants’ confidence that they can perform the behavior required to participate in the intervention) (2). Response options were structured using a 4-point Likert scale. Negatively worded questions were reverse-coded before generating summative scores, and any missing data (3.4%) were imputed as the mean of other items in the measure. The scores were standardized to range from 0 to 100, with higher scores indicating higher acceptability. Internal consistency reliability for acceptability within this sample was Cronbach’s α = 0.84.

### Covariates

Participants self-reported sociodemographic characteristics, including age, gender, race and ethnicity, educational attainment, neighborhood of residence, employment status, receipt of public income assistance, preferred language, level of English proficiency, housing status, social support, medical insurance status, food insecurity,^56^ and relationship status. They also shared their pregnancy status (pregnant or postpartum, or family member of pregnant or postpartum person), parity, history of preterm birth, history of pregnancy loss (e.g., miscarriage, induced abortion, or stillbirth), and any prenatal care attendance during current/recent pregnancy (if pregnant/postpartum participant). Experiences of discrimination were assessed during prenatal care encounters and daily activities, using questions adapted from the Discrimination in Medical Settings Scale and the Everyday Discrimination Scale, respectively.^57,58^ (*Everyday Discrimination Scale*, https://scholar.harvard.edu/davidrwilliams/node/32397).

### Analyses

#### Quantitative

We performed descriptive statistics to examine participants’ sociodemographic characteristics and accessibility and acceptability scores. Prior to bivariate analysis, missing responses to the experience of everyday discrimination and experience of discrimination during prenatal care encounters questions were recoded to the middle response (“Sometimes”). We examined bivariate associations between accessibility and acceptability, as well as the sociodemographic characteristics, pregnancy, obstetric factors, and experiences of discrimination, using linear regression to identify statistically significant associations. Statistically significant predictors (p<0.05) were considered for inclusion in multivariate models. Multivariate models were refined by excluding closely related variables identified through collinearity tests and model fit assessment. Given that some participants answered more than once, we estimated linear mixed-effects models to account for clustering. We performed sensitivity analyses of the above analyses in which we included data only from the first response of individuals who participated in the survey more than once. STATA (version 14) was used to conduct all analyses (StataCorp, *Stata Statistical Software: Release 14*, 2017).

#### Qualitative

We used a framework analysis approach to analyze the qualitative data.^59^ The qualitative lead (OJO) developed an initial deductive codebook based on the interview guide. Six analysts (OJO, PD, KV, EK, HS, and KM) independently coded transcripts using Dedoose software, (SocioCultural Research Consultants, LLC, *Dedoose Version 9.0.107*, 2023, https://www.dedoose.com) and two transcripts were reviewed collaboratively to ensure inter-rater reliability. We compared and discussed coding in detail, refining the codebook, and adding inductive codes. The remaining transcripts were coded individually by balanced pairs of analysts, and the qualitative lead reviewed the coding to ensure consistency. Codes related to the acceptability of the PV model and accessibility of PV services were then queried and evaluated. Analytic summaries for the codes were compiled in a framework table as follows: (1) anticipated affective attitude; (2) experienced affective attitude; (3) self-efficacy; (4) perceived effectiveness; (5) usefulness of PV services; (6) appropriateness of PV services; (7) barriers to access; (8) facilitators of access; and (9) service recommendations and suggestions for improvement.

### Reflexivity

Reflexivity was an integral component of our methodology, particularly in light of co-authors’ varying degrees of involvement in the PV intervention, including development, implementation, and evaluation (MAN, GJ, and KC). Notably, some co-authors also served as community implementing partners (GJ and KC), contributing their critical lived and professional experiences that shaped the design and implementation of PV, as well as the dissemination of key findings. While these dual roles provide important contextual insights, they can also introduce the potential for bias. To address this, we engaged in ongoing reflexive practice to explore how our positionality and individual experiences could affect data collection, analysis, and interpretation. Further, quantitative data were self-administered, and qualitative data were collected and analyzed by team members who were not involved in the implementation, reducing the likelihood of social desirability bias. With respect to qualitative analysis, we employed a systematic approach, including collaborative coding, joint reviews of transcripts, and regular analytic discussions to ensure consistency and challenge our assumptions. While our proximity to the intervention may have shaped our analytical lens, we also consider it a strength when critical reflection and rigorous methodology are maintained.

### Mixed-methods integration

Integration occurred during the design and sampling stages, as described above. We employed a convergent design, aligning questions in the in-depth interview guide with the same framework used in the survey questions to expand upon or explain the quantitative findings. Additionally, the qualitative sample was drawn from the quantitative sample. We also present the quantitative and qualitative findings together in the results section for joint interpretation.

### Ethics approval

This study was conducted in accordance with all relevant ethical regulations, including the Declaration of Helsinki. We obtained ethics approval from the Institutional Review Board of the University of California San Francisco (#20–32393). Verbal informed consent was obtained at PV events after being given information about the study and before survey administration.

## Data availability

The datasets generated and/or analyzed during the current study are not publicly available due to privacy and ethical restrictions but are available from the corresponding author upon reasonable request.

## Coding availability

The underlying code for this study is available upon reasonable request from the corresponding author.

## Acknowledgments

This study was funded by the California Preterm Birth Initiative (A133134) and the California Health Care Foundation (A139605). The funder played no role in the study design, data collection, analysis, and interpretation of data, or the writing of this manuscript. We would like to thank all team members, including Dr. Hannah Sans and Ms. Kobi Miller, for their assistance with qualitative data analysis.

## Author contributions

OJO: Investigation, validation, methodology, data curation, formal analysis, visualization, project administration, writing – original draft, writing – review and editing; AME and AJB: Conceptualization, investigation, validation, methodology, data curation, project administration, writing – original draft, writing – review and editing; MAN: Conceptualization, investigation, validation, methodology, project administration, resources, funding acquisition, writing – original draft, writing – review and editing; KV: Data curation, formal analysis, writing – original draft, writing – review and editing; PCD and EK: formal analysis, writing – original draft, writing – review and editing; CO and JV: writing – original draft, writing – review and editing; NK: data curation, writing – review and editing; GJ and KC: writing – review and editing; PAA: Conceptualization, investigation, validation, methodology, project administration, resources, funding acquisition, supervision, writing – original draft, writing – review and editing.

## Competing interests

All authors declare no financial or non-financial competing interests.

## Notes

### Competing Interest Statement

The authors have declared no competing interest.

### Author Declarations

We obtained ethics approval from the Institutional Review Board of the University of California San Francisco(#2032393).

