## Supplemental Tables for "Accessibility and acceptability of a multi-sector collaborative project to address birth inequities: A mixed-methods study"

Supplementary Table 1. Bivariate sub-group analysis of sociodemographic characteristics, obstetric history, and care discrimination experiences on the accessibility score

|  | Pregnant/Postpartum | | | | | | Family | | | | | |
| --- | --- | --- | --- | --- | --- | --- | --- | --- | --- | --- | --- | --- |
| **Predictor variables** | Cross Tabs | | OLS Regression | | | | Cross Tabs | | OLS Regression | | | |
|  | *Mean* | *SD* | *Coeff.* | *[95% CI]* | | *P-value* | *Mean* | *SD* | *Coeff.* | *[95% CI]* | | *P-value* |
| *Age* |  |  |  |  |  |  |  |  |  |  |  |  |
| 15 - 24 [Reference Group] | 80.0 | 13.1 | 0.0 |  |  |  | 66.7 | 28.9 | 0.0 |  |  |  |
| 25 -34 | 73.5 | 27.5 | -6.5 | -23.8 | 10.8 | 0.247 | 63.0 | 28.6 | -3.7 | -51.5 | 44.0 | 0.821 |
| 35 - 44 | 71.1 | 27.1 | -8.9 | -47.4 | 29.5 | 0.422 | 83.3 | 10.5 | 16.7 | 1.7 | 31.7 | 0.038 |
| 45 and older | 50.0 | 0.0 | -30.0 | -30.0 | -30.0 | 0.000 | 82.8 | 15.1 | 16.1 | -14.2 | 46.4 | 0.189 |
| Unknown | 70.8 | 16.0 | -9.2 | -30.5 | 12.2 | 0.206 | 83.3 | 8.3 | 16.7 | -13.3 | 46.6 | 0.175 |
| *Gender* |  |  |  |  |  |  |  |  |  |  |  |  |
| Female [Reference Group] | 73.1 | 25.0 | 0.0 |  |  |  | 80.2 | 16.0 | 0.0 |  |  |  |
| Male |  |  | … | … | … | … | 66.7 | 35.0 | -13.5 | -46.1 | 19.0 | 0.277 |
| Other/unknown/prefer not to answer | 75.0 | 16.7 | 1.9 | -14.4 | 18.2 | 0.662 | 83.3 | 0.0 | 3.1 | -0.9 | 7.1 | 0.09 |
| *Race/ethnicity* |  |  |  |  |  |  |  |  |  |  |  |  |
| Non-Hispanic black [Reference Group] | 76.7 | 27.3 | 0.0 |  |  |  | 78.6 | 17.6 | 0.0 |  |  |  |
| Hispanic/Latine | 75.3 | 24.6 | -1.4 | -26.7 | 24.0 | 0.839 | 74.4 | 24.3 | -4.2 | -13.1 | 4.7 | 0.232 |
| Multiracial | 69.7 | 19.5 | -7.0 | -30.0 | 16.1 | 0.323 | 88.1 | 8.1 | 9.4 | 2.8 | 16.1 | 0.021 |
| Other/unknown/prefer not to answer | 50.0 | 16.7 | -26.7 | -31.8 | -21.6 | 0.002 | 83.3 | 0.0 | 4.7 | -2.0 | 11.4 | 0.112 |
| *Language* |  |  |  |  |  |  |  |  |  |  |  |  |
| English [Reference Group] | 75.0 | 23.1 | 0.0 |  |  |  | 80.3 | 16.6 | 0.0 |  |  |  |
| Spanish | 72.7 | 26.0 | -2.3 | -31.3 | 26.7 | 0.768 | 73.1 | 25.9 | -7.3 | -15.6 | 1.1 | 0.069 |
| Other/unknown/prefer not to answer | 62.5 | 28.5 | -12.5 | -48.9 | 23.9 | 0.277 | 83.3 | 0.0 | 3.0 | -1.1 | 7.1 | 0.105 |
| *English proficiency* |  |  |  |  |  |  |  |  |  |  |  |  |
| Very well or well [Reference Group] | 75.0 | 23.0 | 0.0 |  |  |  | 79.4 | 19.1 | 0.0 |  |  |  |
| With difficulty | 77.3 | 21.4 | 2.3 | -21.3 | 25.9 | 0.719 | 81.5 | 19.4 | 2.1 | -18.0 | 22.2 | 0.760 |
| Unknown/prefer not to answer | 50.0 | 33.3 | -25.0 | -59.7 | 9.7 | 0.09 | 70.0 | 13.9 | -9.4 | -24.2 | 5.4 | 0.138 |
| *Number of births* |  |  |  |  |  |  |  |  |  |  |  |  |
| None [Reference Group] | 73.3 | 20.7 | 0.0 |  |  |  | 75.8 | 18.8 | 0.0 |  |  |  |
| 1 to 2 | 75.9 | 19.8 | 2.6 | -18.5 | 23.7 | 0.65 | 81.9 | 15.8 | 6.1 | -2.5 | 14.8 | 0.110 |
| 3+ | 67.9 | 34.9 | -5.5 | -67.7 | 56.8 | 0.741 | 81.0 | 14.4 | 5.2 | -13.9 | 24.3 | 0.450 |
| Unknown/prefer not to answer |  |  |  |  |  |  | 70.8 | 30.5 | -4.9 | -49.1 | 39.3 | 0.746 |
| *Prenatal care attendance* |  |  |  |  |  |  |  |  |  |  |  |  |
| No [Reference Group] | 87.5 | 8.3 | 0.0 |  |  |  | … | … | ... |  |  |  |
| Yes | 72.2 | 24.9 | -15.3 | -38.2 | 7.6 | 0.103 | … | … | … | … | … | … |
| Unknown/prefer not to answer | 70.8 | 28.5 | -16.7 | -24.6 | -8.8 | 0.012 | … | … | … | … | … | … |
| *Number of preterm birth pregnancies* |  |  |  |  |  |  |  |  |  |  |  |  |
| None [Reference Group] | 72.5 | 24.6 | 0.0 |  |  |  | 81.4 | 15.2 | 0.0 |  |  |  |
| At least 1 preterm birth | 75.9 | 25.2 | 3.5 | -20.9 | 27.8 | 0.603 | 77.4 | 18.0 | -4.0 | -9.8 | 1.9 | 0.118 |
| Unknown/prefer not to answer/not applicable | 83.3 | 0.0 | 10.9 | -1.6 | 23.4 | 0.065 | 70.8 | 30.5 | -10.5 | -48.9 | 27.8 | 0.446 |
| *Pregnancy loss history* |  |  |  |  |  |  |  |  |  |  |  |  |
| None [Reference Group] | 73.2 | 25.0 | 0.0 |  |  |  | 79.2 | 16.6 | 0.0 |  |  |  |
| Prior pregnancy loss | 73.3 | 25.6 | 0.1 | -27.8 | 28.0 | 0.989 | 83.3 | 14.4 | 4.2 | -1.6 | 9.9 | 0.106 |
| Unknown/prefer not to answer/ not applicable | 72.2 | 9.6 | -1.0 | -23.2 | 21.2 | 0.863 | 74.2 | 27.2 | -4.9 | -42.0 | 32.2 | 0.701 |
| *Education* |  |  |  |  |  |  |  |  |  |  |  |  |
| Less than high school degree [Reference Group] | 77.8 | 20.4 | 0.0 |  |  |  | 69.4 | 26.4 | 0.0 |  |  |  |
| High school graduate, GED, or equivalent | 70.6 | 26.0 | -7.2 | -29.2 | 14.8 | 0.295 | 71.8 | 18.5 | 2.4 | -2.6 | 7.3 | 0.228 |
| Some college, junior college, or vocational school | 86.1 | 13.9 | 8.3 | -31.9 | 48.6 | 0.467 | 84.2 | 11.4 | 14.7 | 8.4 | 21.0 | 0.005 |
| College graduate and professional or graduate school | 58.3 | 29.7 | -19.4 | -28.5 | -10.4 | 0.011 | 89.6 | 12.4 | 20.1 | 11.4 | 28.9 | 0.005 |
| Unknown/prefer not to answer | 77.8 | 17.2 | 0.0 | -45.5 | 45.5 | 1.000 | 83.3 | 16.7 | 13.9 | 5.1 | 22.6 | 0.015 |
| *Employment status* |  |  |  |  |  |  |  |  |  |  |  |  |
| Unemployed [Reference Group] | 76.6 | 22.3 | 0.0 |  |  |  | 80.2 | 17.0 | 0.0 |  |  |  |
| Full-time | 59.1 | 28.2 | -17.5 | -32.8 | -2.1 | 0.039 | 70.0 | 32.1 | -10.2 | -26.8 | 6.5 | 0.148 |
| Part-time | 79.6 | 21.7 | 3.1 | -33.2 | 39.3 | 0.751 | 66.7 | 28.9 | -13.5 | -45.4 | 18.4 | 0.271 |
| Unknown/prefer not to answer | 70.8 | 28.5 | -5.7 | -21.9 | 10.4 | 0.266 | 83.3 | 10.5 | 3.2 | -1.3 | 7.6 | 0.109 |
| *Receiving public assistance* |  |  |  |  |  |  |  |  |  |  |  |  |
| No [Reference Group] | 64.0 | 23.1 | 0.0 |  |  |  | 78.5 | 20.7 | 0.0 |  |  |  |
| Yes | 79.6 | 21.0 | 15.5 | 11.4 | 19.7 | 0.004 | 79.2 | 16.5 | 0.7 | -12.4 | 13.8 | 0.881 |
| Unknown/ prefer not to answer | 69.4 | 37.1 | 5.4 | -39.9 | 50.7 | 0.658 | 83.3 | 0.0 | 4.8 | -0.2 | 9.9 | 0.056 |
| *Residence* |  |  |  |  |  |  |  |  |  |  |  |  |
| Bayview-Hunter's Point (San Francisco) [Reference Group] | 87.1 | 17.0 | 0.0 |  |  |  | 89.2 | 13.1 | 0.0 |  |  |  |
| Other San Francisco | 70.7 | 18.2 | -16.5 | -20.8 | -12.1 | 0.004 | 73.5 | 20.8 | -15.8 | -30.2 | -1.3 | 0.041 |
| East Bay | 47.6 | 26.2 | -39.5 | -82.3 | 3.3 | 0.058 | 70.8 | 16.0 | -18.4 | -31.9 | -4.8 | 0.023 |
| Other/unknown/prefer not to answer | 41.7 | 58.9 | -45.5 | -200.5 | 109.6 | 0.334 | 79.2 | 14.8 | -10.0 | -40.6 | 20.5 | 0.372 |
| *Housing status* |  |  |  |  |  |  |  |  |  |  |  |  |
| Homeless shelter [Reference Group] | 83.3 | 19.2 | 0.0 |  |  |  | 73.3 | 22.4 | 0.0 |  |  |  |
| Owns home or apartment | 72.2 | 34.7 | -11.1 | -11.1 | -11.1 | 0.000 | 93.3 | 9.1 | 20.0 | 9.2 | 30.8 | 0.010 |
| Rents home or apartment | 71.7 | 27.0 | -11.7 | -32.2 | 8.8 | 0.134 | 76.8 | 19.4 | 3.5 | -13.1 | 20.0 | 0.554 |
| Public housing | 76.2 | 23.3 | -7.1 | -41.4 | 27.1 | 0.464 | 85.0 | 16.6 | 11.7 | -1.8 | 25.2 | 0.071 |
| Unknown/Other (e.g., living with someone for free, no living place, transitional housing etc.) | 68.5 | 17.6 | -14.8 | -36.9 | 7.3 | 0.102 | 72.9 | 17.7 | -0.4 | -8.0 | 7.1 | 0.872 |
| *Support* |  |  |  |  |  |  |  |  |  |  |  |  |
| No, not at all [Reference Group] | 91.7 | 9.6 | 0.0 |  |  |  | 77.3 | 11.2 | 0.0 |  |  |  |
| A little | 68.8 | 20.8 | -22.9 | -48.9 | 3.0 | 0.063 | 72.2 | 34.7 | -5.1 | -35.0 | 24.9 | 0.628 |
| Somewhat | 66.7 | 29.7 | -25.0 | -34.8 | -15.2 | 0.008 | 75.9 | 31.3 | -1.3 | -15.8 | 13.1 | 0.787 |
| Yes, definitely | 74.7 | 23.5 | -16.9 | -33.4 | -0.4 | 0.048 | 80.7 | 15.3 | 3.5 | -14.9 | 21.9 | 0.592 |
| Unknown/prefer not to answer |  |  |  |  |  |  | 83.3 | 0.0 | 6.1 | -5.7 | 17.9 | 0.201 |
| *Medical Insurance* |  |  |  |  |  |  |  |  |  |  |  |  |
| Public insurance (e.g., Medicaid, Medi-Cal, etc.) [Reference Group] | 80.3 | 16.8 | 0.0 |  |  |  | 82.5 | 15.3 | 0.0 |  |  |  |
| Private or employer-provided insurance | 60.6 | 30.1 | -19.7 | -36.8 | -2.5 | 0.039 | 81.0 | 6.3 | -1.5 | -12.9 | 9.9 | 0.699 |
| No insurance | 33.3 | 0.0 | -46.9 | -65.0 | -28.8 | 0.008 | 41.7 | 21.5 | -40.8 | -47.0 | -34.6 | 0.000 |
| Unknown/prefer not to answer | 58.3 | 36.1 | -21.9 | -41.1 | -2.8 | 0.039 | 77.8 | 22.8 | -4.7 | -24.0 | 14.6 | 0.495 |
| *Food insecurity (Worried food would run out)* |  |  |  |  |  |  |  |  |  |  |  |  |
| Often true [Reference Group] | 83.3 | 11.8 | 0.0 |  |  |  | 77.1 | 8.6 | 0.0 |  |  |  |
| Sometimes true | 77.2 | 19.4 | -6.1 | -25.5 | 13.2 | 0.305 | 74.1 | 23.7 | -3.0 | -21.7 | 15.7 | 0.644 |
| Never true | 72.2 | 28.1 | -11.1 | -25.3 | 3.1 | 0.078 | 82.4 | 15.6 | 5.3 | -1.3 | 11.9 | 0.084 |
| Unknown/prefer not to answer | 63.6 | 27.7 | -19.7 | -35.9 | -3.4 | 0.035 | 62.5 | 37.0 | -14.6 | -42.4 | 13.3 | 0.194 |
| *Food insecurity (Insufficient funds)* |  |  |  |  |  |  |  |  |  |  |  |  |
| Often true [Reference Group] | 83.3 | 13.6 | 0.0 |  |  |  | 77.8 | 17.2 | 0.0 |  |  |  |
| Sometimes true | 75.4 | 25.1 | -7.9 | -13.5 | -2.4 | 0.026 | 72.7 | 21.4 | -5.1 | -44.0 | 33.9 | 0.707 |
| Never true | 70.2 | 24.6 | -13.2 | -32.2 | 5.9 | 0.097 | 83.9 | 13.8 | 6.1 | -14.0 | 26.2 | 0.405 |
| Unknown/prefer not to answer | 66.7 | 28.9 | -16.7 | -33.3 | -0.1 | 0.05 | 68.8 | 27.4 | -9.0 | -35.6 | 17.5 | 0.358 |
| *Relationship status* |  |  |  |  |  |  |  |  |  |  |  |  |
| Married/partnered, living together [Reference Group] | 72.2 | 26.3 | 0.0 |  |  |  | 65.4 | 26.8 | 0.0 |  |  |  |
| Married/partnered, not living together | 73.3 | 16.1 | 1.1 | -21.1 | 23.4 | 0.850 | 77.8 | 17.2 | 12.4 | -1.0 | 25.8 | 0.060 |
| Single | 82.1 | 19.0 | 9.9 | -26.2 | 46.1 | 0.359 | 83.3 | 12.8 | 17.9 | 6.6 | 29.3 | 0.015 |
| Other/unknown/prefer not to answer | 60.4 | 32.0 | -11.8 | -60.8 | 37.1 | 0.408 | 91.7 | 11.8 | 26.3 | -1.2 | 53.8 | 0.056 |
| *Everyday discrimination* |  |  |  |  |  |  |  |  |  |  |  |  |
| Never [Reference Group] | 78.8 | 16.8 | 0.0 |  |  |  | 68.5 | 29.4 | 0.0 |  |  |  |
| Rarely | 79.2 | 21.5 | 0.4 | -27.3 | 28.0 | 0.958 | 81.0 | 12.8 | 12.4 | -9.1 | 33.9 | 0.163 |
| Sometimes | 75.8 | 26.2 | -3.0 | -9.9 | 4.0 | 0.209 | 78.3 | 18.4 | 9.7 | -6.7 | 26.2 | 0.156 |
| Often | 50.0 | 22.0 | -28.8 | -32.0 | -25.6 | 0.001 | 86.7 | 10.5 | 18.1 | 0.3 | 36.0 | 0.048 |
| *Discrimination during prenatal care encounters* |  |  |  |  |  |  |  |  |  |  |  |  |
| Never [Reference Group] | 81.5 | 18.9 | 0.0 |  |  |  | 80.6 | 16.2 | 0.0 |  |  |  |
| Rarely | 76.7 | 19.7 | -4.8 | -27.8 | 18.1 | 0.462 | 85.7 | 6.3 | 5.1 | -6.0 | 16.1 | 0.241 |
| Sometimes | 64.2 | 28.8 | -17.3 | -55.4 | 20.7 | 0.189 | 66.7 | 25.5 | -14.0 | -43.7 | 15.8 | 0.232 |
| Often | 66.7 | 33.3 | -14.8 | -45.1 | 15.5 | 0.170 | 90.0 | 9.1 | 9.4 | 0.3 | 18.5 | 0.047 |

Abbreviations: 95% CI: 95% confidence interval; SD: standard deviation

Supplementary Table 2. Bivariate sub-group analysis of sociodemographic characteristics, obstetric history, and care discrimination experiences on the acceptability score

|  | Pregnant/Postpartum | | | | | | Family | | | | | |
| --- | --- | --- | --- | --- | --- | --- | --- | --- | --- | --- | --- | --- |
| Predictor variables | Cross Tabs | | OLS Regression | | | | Cross Tabs | | OLS Regression | | | |
|  | Mean | SD | Coeff. | *[95% CI]* | | *P-value* | Mean | SD | Coeff. | *[95% CI]* | | *P-value* |
| *Age* |  |  |  |  |  |  |  |  |  |  |  |  |
| 15 - 24 [Reference Group] | 96.2 | 5.4 | 0.0 |  |  |  | 79.4 | 27.9 | 0.0 |  |  |  |
| 25 -34 | 91.3 | 17.5 | -4.8 | -6.7 | -3.0 | 0.008 | 84.7 | 18.3 | 5.3 | -3.9 | 14.5 | 0.164 |
| 35 - 44 | 88.9 | 16.9 | -7.3 | -28.7 | 14.0 | 0.279 | 95.2 | 8.0 | 15.9 | -12.7 | 44.4 | 0.175 |
| 45 and older | 90.5 | 0.0 | -5.7 | -5.7 | -5.7 | 0.000 | 95.4 | 10.0 | 16.0 | -9.5 | 41.5 | 0.139 |
| Unknown | 79.5 | 17.5 | -16.7 | -59.8 | 26.5 | 0.238 | 99.0 | 3.0 | 19.7 | -1.6 | 41.0 | 0.06 |
| *Gender* |  |  |  |  |  |  |  |  |  |  |  |  |
| Female [Reference Group] | 89.9 | 16.4 |  |  |  |  | 93.7 | 12.2 | 0.0 |  |  |  |
| Male |  |  | … | … | … | … | 88.9 | 19.4 | -4.8 | -17.7 | 8.2 | 0.327 |
| Other/unknown/prefer not to answer | 94.8 | 7.6 | 4.9 | 1.6 | 8.2 | 0.024 | 100.0 | 0.0 | 6.3 | 0.6 | 12.1 | 0.038 |
| *Race/ethnicity* |  |  |  |  |  |  |  |  |  |  |  |  |
| Non-Hispanic black [Reference Group] | 93.3 | 13.9 | 0.0 |  |  |  | 94.6 | 12.4 | 0.0 |  |  |  |
| Hispanic/Latine | 86.5 | 18.9 | -6.8 | -36.6 | 23.0 | 0.430 | 91.1 | 14.0 | -3.5 | -10.4 | 3.3 | 0.200 |
| Multiracial | 93.5 | 10.5 | 0.2 | -14.8 | 15.2 | 0.965 | 90.5 | 14.8 | -4.2 | -10.8 | 2.4 | 0.138 |
| Other/unknown/prefer not to answer | 95.8 | 8.3 | 2.5 | -15.5 | 20.5 | 0.611 | 100.0 | 0.0 | 5.4 | -1.2 | 11.9 | 0.081 |
| *Language* |  |  |  |  |  |  |  |  |  |  |  |  |
| English [Reference Group] | 94.9 | 11.2 | 0.0 |  |  |  | 93.9 | 12.7 | 0.0 |  |  |  |
| Spanish | 83.7 | 19.8 | -11.2 | -33.9 | 11.5 | 0.167 | 90.5 | 14.9 | -3.4 | -12.5 | 5.7 | 0.318 |
| Other/unknown/prefer not to answer | 91.9 | 11.4 | -3.0 | -21.5 | 15.5 | 0.555 | 98.1 | 4.3 | 4.2 | -2.4 | 10.8 | 0.136 |
| *English proficiency* |  |  |  |  |  |  |  |  |  |  |  |  |
| Very well or well [Reference Group] | 94.6 | 10.5 | 0.0 |  |  |  | 93.2 | 13.9 | 0.0 |  |  |  |
| With difficulty | 85.1 | 16.8 | -9.5 | -35.7 | 16.7 | 0.259 | 93.7 | 11.4 | 0.5 | -11.7 | 12.6 | 0.913 |
| Unknown/prefer not to answer | 71.0 | 27.0 | -23.6 | -28.2 | -19.0 | 0.002 | 95.2 | 5.2 | 2.0 | -4.3 | 8.4 | 0.382 |
| *Number of births* |  |  |  |  |  |  |  |  |  |  |  |  |
| None [Reference Group] | 92.4 | 13.3 | 0.0 |  |  |  | 90.5 | 17.6 | 0.0 |  |  |  |
| 1 to 2 | 95.2 | 8.9 | 2.9 | -10.8 | 16.5 | 0.463 | 94.4 | 10.9 | 3.9 | -18.5 | 26.3 | 0.615 |
| 3+ | 79.1 | 23.2 | -13.2 | -69.0 | 42.5 | 0.415 | 94.9 | 9.4 | 4.4 | -19.8 | 28.6 | 0.602 |
| Unknown/prefer not to answer | 83.3 | 0.0 | -9.0 | -29.5 | 11.4 | 0.197 | 92.6 | 16.3 | 2.1 | -27.6 | 31.8 | 0.835 |
| *Prenatal care attendance* |  |  |  |  |  |  |  |  |  |  |  |  |
| No [Reference Group] | 98.8 | 2.4 | 0.0 |  |  |  | … | … | … |  |  |  |
| Yes | 89.0 | 16.8 | -9.9 | -16.3 | -3.4 | 0.022 | … | … | … | … | … | … |
| Unknown/prefer not to answer | 96.7 | 7.5 | -2.1 | -4.7 | 0.4 | 0.07 | … | … | … | … | … | … |
| *Number of preterm birth pregnancies* |  |  |  |  |  |  |  |  |  |  |  |  |
| None [Reference Group] | 92.0 | 14.5 | 0.0 |  |  |  | 95.9 | 9.8 | 0.0 |  |  |  |
| At least 1 preterm birth | 82.0 | 21.8 | -10.0 | -34.5 | 14.6 | 0.222 | 88.1 | 15.7 | -7.8 | -19.1 | 3.4 | 0.113 |
| Unknown/prefer not to answer/not applicable | 89.3 | 8.4 | -2.7 | -9.3 | 3.8 | 0.218 | 92.6 | 16.3 | -3.3 | -19.3 | 12.6 | 0.553 |
| *Pregnancy loss history* |  |  |  |  |  |  |  |  |  |  |  |  |
| None [Reference Group] | 91.1 | 16.0 | 0.0 |  |  |  | 94.7 | 11.7 | 0.0 |  |  |  |
| Prior pregnancy loss | 90.9 | 15.4 | -0.2 | -22.9 | 22.6 | 0.976 | 94.7 | 11.0 | 0.0 | -7.4 | 7.4 | 1.000 |
| Unknown/prefer not to answer/ not applicable | 81.5 | 17.9 | -9.5 | -24.9 | 5.9 | 0.117 | 88.9 | 16.8 | -5.8 | -30.8 | 19.1 | 0.512 |
| *Education* |  |  |  |  |  |  |  |  |  |  |  |  |
| Less than high school degree [Reference Group] | 88.7 | 10.9 | 0.0 |  |  |  | 86.5 | 19.6 | 0.0 |  |  |  |
| High school graduate, GED, or equivalent | 89.1 | 19.5 | 0.4 | -18.6 | 19.3 | 0.942 | 93.8 | 10.7 | 7.3 | -6.3 | 20.8 | 0.186 |
| Some college, junior college, or vocational school | 100.0 | 0.0 | 11.3 | -4.1 | 26.7 | 0.087 | 95.7 | 9.5 | 9.2 | -7.1 | 25.5 | 0.170 |
| College graduate and professional or graduate school | 86.5 | 18.6 | -2.2 | -25.5 | 21.1 | 0.723 | 95.8 | 11.8 | 9.3 | 1.1 | 17.5 | 0.036 |
| Unknown/prefer not to answer | 85.4 | 16.5 | -3.3 | -20.3 | 13.6 | 0.486 | 97.6 | 4.8 | 11.1 | -2.2 | 24.4 | 0.077 |
| *Employment status* |  |  |  |  |  |  |  |  |  |  |  |  |
| Unemployed [Reference Group] | 88.3 | 17.3 | 0.0 |  |  |  | 93.9 | 11.6 | 0.0 |  |  |  |
| Full-time | 92.6 | 9.6 | 4.3 | -12.0 | 20.6 | 0.375 | 83.8 | 22.7 | -10.1 | 0.2 | -31.7 | 0.235 |
| Part-time | 98.4 | 3.4 | 10.1 | -5.1 | 25.2 | 0.103 | 100.0 | 0.0 | 6.1 | 0.0 | 0.2 | 0.047 |
| Unknown/prefer not to answer | 83.3 | 26.2 | -5.0 | -16.7 | 6.7 | 0.208 | 95.2 | 12.6 | 1.4 | 0.8 | -16.9 | 0.828 |
| *Receiving public assistance* |  |  |  |  |  |  |  |  |  |  |  |  |
| No [Reference Group] | 91.4 | 10.6 | 0.0 |  |  |  | 92.0 | 15.2 | 0.0 |  |  |  |
| Yes | 90.9 | 16.1 | -0.5 | -10.8 | 9.9 | 0.867 | 94.8 | 9.3 | 2.8 | -7.8 | 13.5 | 0.460 |
| Unknown/ prefer not to answer | 84.7 | 25.7 | -6.7 | -25.8 | 12.4 | 0.270 | 100.0 | 0.0 | 8.0 | 0.0 | 15.9 | 0.05 |
| *Residence* |  |  |  |  |  |  |  |  |  |  |  |  |
| Bayview-Hunter's Point (San Francisco) [Reference Group] | 91.5 | 12.3 | 0.0 |  |  |  | 95.8 | 6.0 | 0.0 |  |  |  |
| Other San Francisco | 91.6 | 16.0 | 0.1 | -7.6 | 7.8 | 0.948 | 92.4 | 14.1 | -3.4 | -14.3 | 7.5 | 0.395 |
| East Bay | 92.5 | 10.9 | 1.0 | -17.0 | 19.1 | 0.829 | 89.3 | 15.7 | -6.5 | -10.3 | -2.7 | 0.012 |
| Other/unknown/prefer not to answer | 65.9 | 32.3 | -25.6 | -75.5 | 24.3 | 0.158 | 94.2 | 17.5 | -1.6 | -26.6 | 23.4 | 0.85 |
| *Housing status* |  |  |  |  |  |  |  |  |  |  |  |  |
| Homeless shelter [Reference Group] | 93.9 | 9.0 | 0.0 |  |  |  | 91.4 | 14.4 | 0.0 |  |  |  |
| Owns home or apartment | 100.0 | 0.0 | 6.1 | 0.1 | 12.1 | 0.048 | 99.0 | 2.1 | 7.6 | -1.6 | 16.9 | 0.079 |
| Rents home or apartment | 91.5 | 15.0 | -2.3 | -9.3 | 4.6 | 0.284 | 94.6 | 12.4 | 3.1 | -8.1 | 14.4 | 0.440 |
| Public housing | 79.6 | 24.7 | -14.3 | -65.4 | 36.8 | 0.352 | 91.9 | 12.5 | 0.5 | -0.8 | 1.8 | 0.323 |
| Unknown/Other (e.g., living with someone for free, no living place, transitional housing etc.) | 88.8 | 15.1 | -5.1 | -28.5 | 18.4 | 0.451 | 89.9 | 17.2 | -1.5 | -21.7 | 18.8 | 0.831 |
| *Support* |  |  |  |  |  |  |  |  |  |  |  |  |
| No, not at all [Reference Group] | 91.7 | 13.7 | 0.0 |  |  |  | 86.1 | 17.6 | 0.0 |  |  |  |
| A little | 82.7 | 24.3 | -8.9 | -47.0 | 29.1 | 0.419 | 100.0 | 0.0 | 13.9 | -5.8 | 33.5 | 0.111 |
| Somewhat | 90.5 | 12.7 | -1.2 | -39.7 | 37.3 | 0.906 | 85.2 | 18.5 | -1.0 | -24.4 | 22.4 | 0.904 |
| Yes, definitely | 92.3 | 15.0 | 0.6 | -30.6 | 31.8 | 0.941 | 97.3 | 6.7 | 11.2 | -9.2 | 31.5 | 0.179 |
| Unknown/prefer not to answer | 83.3 | 0.0 | -8.3 | -40.6 | 23.9 | 0.382 | 100.0 | 0.0 | 13.9 | -5.8 | 33.5 | 0.111 |
| *Medical Insurance* |  |  |  |  |  |  |  |  |  |  |  |  |
| Public insurance (e.g., Medicaid, Medi-Cal, etc.) [Reference Group] | 94.8 | 9.4 | 0.0 |  |  |  | 95.4 | 11.0 | 0.0 |  |  |  |
| Private or employer-provided insurance | 86.1 | 14.4 | -8.7 | -35.2 | 17.9 | 0.295 | 95.2 | 12.6 | -0.1 | -10.8 | 10.6 | 0.973 |
| No insurance | 61.9 | 0.0 | -32.9 | -40.5 | -25.3 | 0.003 | 81.0 | 20.6 | -14.4 | -18.1 | -10.7 | 0.001 |
| Unknown/prefer not to answer | 76.5 | 30.4 | -18.3 | -33.1 | -3.5 | 0.033 | 88.4 | 15.3 | -6.9 | -14.3 | 0.5 | 0.059 |
| *Food insecurity (Worried food would run out)* |  |  |  |  |  |  |  |  |  |  |  |  |
| Often true [Reference Group] | 93.3 | 12.4 | 0.0 |  |  |  | 88.1 | 14.8 | 0.0 |  |  |  |
| Sometimes true | 94.4 | 9.6 | 1.1 | -15.8 | 17.9 | 0.810 | 92.6 | 11.2 | 4.5 | -6.5 | 15.5 | 0.284 |
| Never true | 95.0 | 7.9 | 1.7 | -10.5 | 13.9 | 0.614 | 95.6 | 11.3 | 7.6 | 2.7 | 12.4 | 0.016 |
| Unknown/prefer not to answer | 74.4 | 24.6 | -18.9 | -35.6 | -2.2 | 0.039 | 88.6 | 20.6 | 0.5 | -20.7 | 21.6 | 0.947 |
| *Food insecurity (Insufficient funds)* |  |  |  |  |  |  |  |  |  |  |  |  |
| Often true [Reference Group] | 93.2 | 10.9 | 0.0 |  |  |  | 87.3 | 16.4 | 0.0 |  |  |  |
| Sometimes true | 90.6 | 13.8 | -2.6 | -28.8 | 23.6 | 0.714 | 92.2 | 10.9 | 4.9 | -18.8 | 28.6 | 0.557 |
| Never true | 91.5 | 16.5 | -1.7 | -16.6 | 13.1 | 0.668 | 96.8 | 8.1 | 9.5 | 0.9 | 18.1 | 0.039 |
| Unknown/prefer not to answer | 85.5 | 21.8 | -7.7 | -29.1 | 13.7 | 0.261 | 87.8 | 21.7 | 0.5 | -20.7 | 21.7 | 0.942 |
| *Relationship status* |  |  |  |  |  |  |  |  |  |  |  |  |
| Married/partnered, living together [Reference Group] | 90.0 | 12.2 | 0.0 |  |  |  | 90.8 | 14.9 | 0.0 |  |  |  |
| Married/partnered, not living together | 93.3 | 14.9 | 3.3 | -24.2 | 30.8 | 0.655 | 91.3 | 21.4 | 0.4 | -28.3 | 29.1 | 0.965 |
| Single | 96.9 | 7.8 | 6.9 | -5.5 | 19.3 | 0.138 | 94.4 | 10.8 | 3.6 | -7.2 | 14.3 | 0.367 |
| Other/unknown/prefer not to answer | 77.5 | 26.5 | -12.5 | -38.2 | 13.2 | 0.172 | 98.4 | 2.7 | 7.6 | 0.5 | 14.7 | 0.042 |
| *Everyday discrimination* |  |  |  |  |  |  |  |  |  |  |  |  |
| Never [Reference Group] | 96.1 | 12.9 | 0.0 |  |  |  | 89.9 | 15.1 | 0.0 |  |  |  |
| Rarely | 91.6 | 8.1 | -4.5 | -9.7 | 0.6 | 0.064 | 95.6 | 13.9 | 5.6 | -15.5 | 26.7 | 0.458 |
| Sometimes | 85.1 | 21.9 | -11.0 | -18.6 | -3.3 | 0.025 | 94.4 | 11.4 | 4.5 | -2.1 | 11.1 | 0.118 |
| Often | 93.1 | 9.8 | -3.0 | -5.9 | 0.0 | 0.049 | 91.4 | 13.2 | 1.5 | -2.8 | 5.8 | 0.355 |
| *Discrimination during prenatal care encounters* |  |  |  |  |  |  |  |  |  |  |  |  |
| Never [Reference Group] | 95.5 | 10.4 | 0.0 |  |  |  | 97.5 | 5.1 | 0.0 |  |  |  |
| Rarely | 89.1 | 16.3 | -6.4 | -18.9 | 6.1 | 0.158 | 92.5 | 10.9 | -5.0 | -12.8 | 2.8 | 0.133 |
| Sometimes | 85.4 | 18.9 | -10.1 | -29.7 | 9.5 | 0.156 | 85.4 | 20.3 | -12.2 | -31.6 | 7.3 | 0.141 |
| Often | 100.0 | 0.0 | 4.5 | -5.5 | 14.5 | 0.194 | 92.4 | 14.5 | -5.2 | -7.5 | -2.8 | 0.006 |

Abbreviations: 95% CI: 95% confidence interval; SD: standard deviation

Supplementary Table 3. Sensitivity analysis of acceptability and accessibility scores

|  |  | Combined Sample | Pregnant/postpartum | Family |
| --- | --- | --- | --- | --- |
| **Acceptability** | *N* | 85 | 45 | 40 |
|  | *Score* | 90.8 | 90.5 | 91.2 |
|  | *SD* | 15.2 | 15.9 | 14.5 |
|  | *Minimum Score* | 28.6 | 28.6 | 47.6 |
|  | *Maximum Score* | 100.0 | 100.0 | 100.0 |
| **Accessibility** | *N* | 87 | 46 | 41 |
|  | *Mean* | 74.5 | 71.0 | 78.5 |
|  | *SD* | 22.6 | 24.2 | 20.2 |
|  | *Minimum Score* | 0.0 | 0.0 | 16.7 |
|  | *Maximum Score* | 100.0 | 100.0 | 100.0 |

Abbreviations: SD: standard deviation

Supplementary Table 4. Accessibility scale items

| **Item No.** | **Variable name** | **Question** | **Recoded response options** |
| --- | --- | --- | --- |
| 1 | Actual time to PV | How long did it take you to get to Pop-Up Village from where you typically stay? |  |
|  |  | 0, More than 1 hour | 0, More than 1 hour |
|  |  | 1, Between 30 minutes - 1 hour | 1, Between 30 minutes - 1 hour |
|  |  | 2, Between 15 - 30 minutes | 1, Between 15-30 minutes |
|  |  | 3, Between 5 - 15 minutes | 2, Between 5-15 minutes |
|  |  | 4, Less than five minutes | 2, Less than five minutes |
| 2 | Perceived time to PV | How do you feel about the amount of time it took you to get to Pop-Up Village? |  |
|  |  | 0, It was too long |  |
|  |  | 1, It wasn't too long |  |
|  |  | 2, It was just right |  |
| 3 | Comparative accessibility | Compared to how you access your usual sources of care and resources, would you say the services at Pop-Up Village are… |  |
|  |  | 0, Harder to access |  |
|  |  | 1, About the same |  |
|  |  | 2, Easier to access |  |
|  |  | 3, Not applicable | 1, About the same |
